# The effect of COVID-19 vaccination and booster on maternal-fetal outcomes: a retrospective multicenter cohort study

**DOI:** 10.1101/2022.08.12.22278727

**Authors:** Samantha N. Piekos, Yeon Mi Hwang, Ryan T. Roper, Tanya Sorensen, Nathan D. Price, Leroy Hood, Jennifer J. Hadlock

**Affiliations:** Institute for Systems Biology, Seattle, WA, USA; Swedish Health Services, Swedish Medical Center, Seattle, WA, USA; Thorne HealthTech, New York, NY, USA

**Keywords:** SARS-CoV-2, COVID-19 vaccination, booster, maternal-fetal health, maternal infection, preterm birth, stillbirth

## Abstract

**Background:** COVID-19 infection in pregnant people has previously been shown to increase the risk for poor maternal-fetal outcomes. Despite this, there has been a lag in COVID-19 vaccination in pregnant people due to concerns over the potential effects of the vaccine on maternal-fetal outcomes. Here we examine the impact of COVID-19 vaccination and booster on maternal COVID-19 breakthrough infections and birth outcomes.

**Methods:** This was a retrospective multicenter cohort study on the impact of COVID-19 vaccination on maternal-fetal outcomes for people that delivered (n=86,833) at Providence St. Joseph Health across Alaska, California, Montana, Oregon, New Mexico, Texas, and Washington from January 26, 2021 through July 11, 2022. Cohorts were defined by vaccination status at time of delivery: unvaccinated (n=48,492), unvaccinated propensity score matched (n=26,790), vaccinated (n=26,792; two doses of mRNA-1273 Moderna or BNT162b2 Pfizer-BioNTech), and/or boosted (n=7,616). The primary outcome was maternal COVID-19 infection. COVID-19 vaccination status at delivery, COVID-19 infection-related health care, preterm birth (PTB), stillbirth, very low birth weight (VLBW), and small for gestational age (SGA) were evaluated as secondary outcomes.

**Findings:** Vaccinated pregnant people were significantly less likely to have a maternal COVID-19 infection than unvaccinated matched (p<0.0001) pregnant people. During a maternal COVID-19 infection, vaccinated pregnant people had similar rates of hospitalization (p=0.23), but lower rates of supplemental oxygen (p<0.05) or vasopressor (p<0.05) use than those in an unvaccinated matched cohort. Compared to an unvaccinated matched cohort, vaccinated people had significantly lower stillbirth rate (p<0.01) as well as no difference in rate of PTB (p=0.35), SGA (p=0.79), or rate of VLBW (>1,500 g; 0.31). Vaccinated people who were boosted had significantly lower rates of maternal COVID-19 infections (p<0.0001), COVID-19 related hospitalization (p<0.05), PTB (p<0.05), stillbirth (p<0.01), SGA (p<0.05), and VLBW (p<0.01), compared to vaccinated people that did not receive a third booster dose five months after completing the initial vaccination series.

**Interpretation:** COVID-19 vaccination protects against adverse maternal-fetal outcomes with booster doses conferring additional protection against COVID-19 infection. It is therefore important for pregnant people to have high priority status for vaccination, and for them to stay current with their COVID-19 vaccination schedule.

**Funding:** This study was funded by the National Institute for Child Health & Human Development and the William O. and K. Carole Ellison Foundation.

## Introduction

Maternal COVID-19 infection is a serious risk for maternal-fetal health. Pregnant people are at increased risk of morbidity and mortality from maternal COVID-19 infections.^1–4^ There is also increased risk for preterm birth (PTB), stillbirth, small for gestational age (SGA), and decreased birth weight following a maternal COVID-19 infection.^5–10^ This makes pregnant people an important population of focus for COVID-19 vaccination to promote maternal-fetal health. However, pregnant people were excluded from all COVID-19 vaccination trials, resulting in a lack of information about COVID-19 vaccination safety and efficacy in this population.^11^ This has led to understandable vaccine hesitancy and reduced vaccine uptake in the pregnant population.^9,12–16^ Pregnant people’s concerns include potential harmful side effects to their developing baby and the lack of safety and efficacy data in pregnant people.^12–14,16^ Further, persistent disparities have been observed in COVID-19 vaccination rates with across race, ethnicity, and socioeconomic status.^9,15,17^ Several initial reports indicate that COVID-19 vaccination in pregnant people protects against COVID-19 infection.^18–20^ Vaccine antibodies can pass through the umbilical cord to the fetus *in utero*, which means that COVID-19 vaccination during pregnancy protects the subsequent neonate.^21–23^ The highest levels of antibodies detected in the maternal and umbilical cord blood follow a third mRNA booster dose during the third trimester of pregnancy,^23^ and there is reduced risk of hospitalization related to COVID-19 in the first six months of life for infants born to mothers who were vaccinated during pregnancy.^24^ Initial reports found no difference in rates of PTB, SGA, stillbirth, perinatal mortality, infant mortality, or neonatal hospitalization rates between people that are vaccinated versus unvaccinated.^9,25–27^ However, previous studies on the impact of COVID-19 vaccination on maternal-fetal outcomes remain limited in their depth, scope, breadth of outcomes evaluated, and diversity of study population. Further, there has not been a large retrospective cohort study that examines the impact of a third mRNA booster dose on maternal-fetal outcomes.

Here, we investigate the impact of COVID-19 vaccination and booster on maternal-fetal outcomes including COVID-19 infection, PTB, stillbirth, and very low birth weight (VLBW). We examined five questions in this study: who receives the COVID-19 mRNA vaccine, what impact does COVID-19 mRNA vaccination have on maternal COVID-19 have on infection rate, what impact does the COVID-19 mRNA vaccination have on COVID-19 severity, what effect does COVID-19 mRNA vaccine have on birth outcomes, and how does third mRNA booster impact maternal-fetal outcomes. We report descriptive and quantitative analyses on vaccination, COVID-19 infection in pregnant people, and differences in the composition of vaccinated and unvaccinated populations. In addition, we examine the severity of maternal COVID-19 in vaccinated and unvaccinated individuals. Finally, we evaluate birth outcomes in unvaccinated, vaccinated, and boosted individuals.

## Methods

### Study setting and participants

Providence St. Joseph Health (PSJH) is an integrated U.S. community healthcare system that provides care in urban and rural settings across seven states: Alaska, California, Montana, Oregon, New Mexico, Texas, and Washington. This retrospective cohort study used clinical data from PSJH electronic health records (EHR) for pregnant patients who delivered at PSJH sites from January 26, 2021 through July 11, 2022. Care related to SARS-CoV-2 infection and delivery was observed.

Vaccination status was determined using PSJH EHR data, which includes imported state vaccination records. Cohorts were determined by vaccination status at delivery as defined in Supplemental Table 1. The main cohorts examined were people that did not have a positive SARS-CoV-2 NAAT test prior to the start of pregnancy and were either unvaccinated, or vaccinated with an mRNA vaccine (2-doses of mRNA-1273 Moderna or BNT162b2 Pfizer-BioNTech; Figure 1). We also considered subsets of the vaccinated population that either, by delivery, had received a third mRNA dose (booster), or had not received a booster when their last COVID-19 vaccine dose was more than five months prior. Cohorts were limited to maternal ages from 18 to 45 years with singleton pregnancies that delivered after 20 weeks gestation. COVID-19 infection was determined by a positive SARS-CoV-2 NAAT test. PSJH tests anyone with COVID-19 symptoms and screens patients two days prior to admission for a planned procedure.

**Figure 1.**
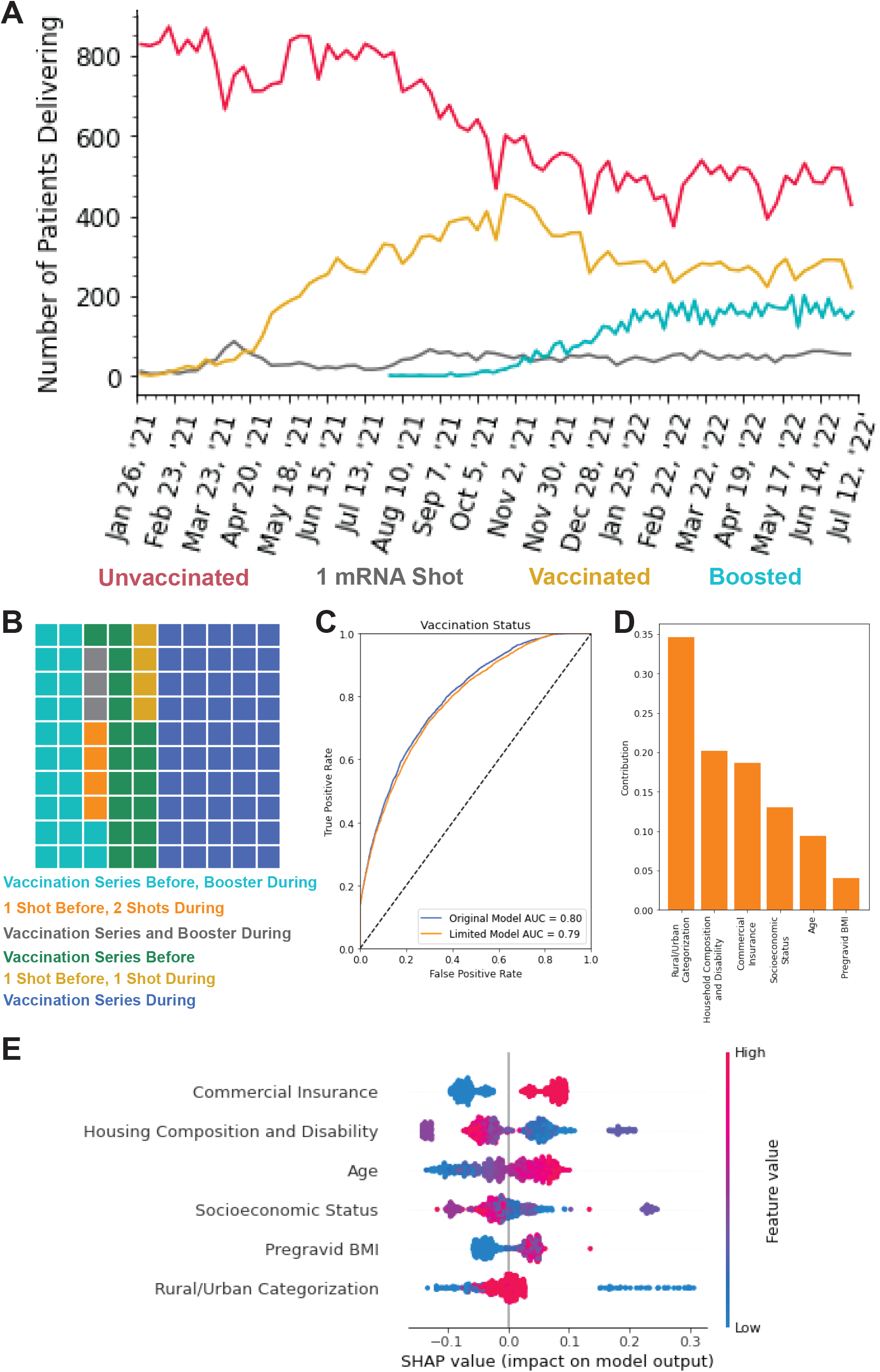
Assessment of characteristics associated with vaccination status. **A:** Weekly counts of vaccination status - unvaccinated (red), one mRNA dose (grey), vaccinated, two mRNA doses only (gold), or boosted (teal) - at delivery for deliveries occurring January 26, 2021 through July 11, 2022. **B:** Waffle plot of the timing of the COVID-19 vaccine doses in relation to pregnancy. **C:** The area under the curve (AUC) of the gradient boosting model predicting vaccination status of pregnant people at delivery based on 20 demographic, comorbidity, and geographical features (blue) or the top six most important features only (orange). **D:** Impurity-based feature importance evaluating the contribution of the six features in the limited gradient boosting model, which predicts vaccination status at delivery. The importance of a feature is computed as the normalized total reduction of the criterion brought by that feature. The higher the value, the more important the feature. **E:** The contribution of the six features in the gradient boosting limited model towards predicting vaccination status at delivery as measured by the Shapley algorithm and reported as the SHAP value. This value is the average marginal contribution of a feature value across all permutations of features providing insight into the degree of influence of the feature on an individual’s predicted vaccination status at delivery. Each line represents a feature, and each dot represents a sample. The dot color represents the value of the feature for the sample, with red being a high value and blue being a low value for that feature across all samples. This evaluation was performed on 1000 patients randomly selected from the test set. SHAP=Shapley additive explanations.

Propensity score matching was used to account for known demographic, lifestyle, geographical, and clinical characteristics associated with negative maternal-fetal outcomes to generate an unvaccinated matched cohort. An unsupervised learning model with k-nearest neighbors (k=1) was used to match with replacement by the propensity logit metric across 20 covariates (Supplemental Table 4) using Python library PsmPy (version 0.2.8).^28^ This identified, for each vaccinated person, the most similar unvaccinated person across 20 variables to create the unvaccinated matched cohort of 26,790 people, which had improved representation of matched covariates with a small (absolute standard mean difference <0.2) effect size for each variable.^29^

This cohort study was reported following STROBE guidelines.^30^ All procedures were reviewed and approved by the Institutional Review Board at the PSJH through expedited review (study number STUDY2020000196). Consent was waived because disclosure of protected health information for the study was determined to involve no more than minimal risk to the privacy of individuals.

### Outcomes

The primary outcome of this study was the rate of COVID-19 infection during pregnancy. A secondary outcome was the timing of breakthrough infection from full vaccination status. Other secondary outcomes reported for maternal COVID-19 infection during Omicron dominance (infection after December 24, 2021) were COVID-19 hospitalization rates, supplemental oxygen use, vasopressor use, COVID-19 severity, number of medications, and number of diagnoses. Additional maternal outcomes reported include rates of gestational diabetes, gestational hypertension, preeclampsia, and severe preeclampsia. Secondary birth outcomes included PTB, stillbirth, SGA (bottom tenth fetal growth percentile at delivery), low birth weight (LBW) (<2,500 g), and VLBW (<1,500 g) rates. Birth weight, gestational age at delivery, and PTB categories were also examined.

Timing of variant dominance was determined by when the variant accounted for >50% of infections as reported by the Center for Disease Control (CDC) genomic surveillance for SARS-CoV-2 in the western United States (Alaska, Idaho, Oregon, and Washington), shown in (Supplemental Table 8).^31,32^ COVID-19 infection severity was determined by patient outcomes in the two weeks prior to and eight weeks following a positive SARS-CoV-2 NAAT test. Mild cases were those that required neither hospitalization nor supplemental oxygen use. Moderate cases were those that required hospitalization and/or low-flow oxygen use. Severe cases required high-flow oxygen use or mechanical ventilation. Death during this period was also reported.

### Statistical analyses and machine learning

Detailed description of the statistical analyses and machine learning applied in this study are in the Supplemental Methods section of Supplemental Materials.

## Results

### Identifying characteristics of vaccinated pregnant people

In people with no recorded COVID-19 infection before the start of pregnancy, we observed 48,492 unvaccinated people and 26,792 people that received two or more doses of either mRNA-1273 Moderna or BNT162b2 Pfizer-BioNTech at least two weeks prior to delivery (Figure 1; Supplemental Table 1). Patients in the vaccinated cohort were more likely to be older, Asian, non-Hispanic, non-smoking or not users of illicit drugs (Table 1, Supplemental Tables 2-3). They are also more likely to have a higher pregravid body mass index (BMI), commercial insurance, no history of PTB, lower parity, lower gravidity, chronic diabetes, chronic hypertension, gestational diabetes, a male fetus, and a cesarean section delivery (Table 1, Supplemental Tables 2-5). Finally, vaccinated people are more likely to live in urban areas and live in areas with lower social vulnerability across all four CDC social vulnerability index (SVI) themes (Table 1, Supplemental Tables 2-5). There was no significant difference in rates of gestational hypertension, preeclampsia, or severe preeclampsia between vaccinated and unvaccinated people (Table 1; Supplemental Table 2-5). During the study period (January 26, 2021 - July 11, 2022), the number of vaccinated or boosted individuals at delivery increases as the number of unvaccinated individuals decline before plateauing at the start of 2022 (Figure 1A). 83.0% (22,226/26,792) of people in the vaccinated cohort received at least one mRNA dose during their pregnancy and 28.4% (7,616/26,792) of the vaccinated cohort received their booster prior to delivery (Figure 1B).

**Table 1.** Demographic, comorbidities, birth characteristics, and geographical features of vaccinated and unvaccinated pregnant people. Distribution of race, ethnicity, pregravid BMI, age, parity, gravidity, fetal sex, mode of delivery, and rural/urban categorization for people vaccinated with mRNA vaccine (n=26,792) or unvaccinated (n=48,492) at delivery. Number of people who have commercial insurance, smoke, use illicit drugs, have previously delivered prematurely, or have common pregnancy-related comorbidities. Median and Interquartile Range (IQR) of CDC Social Vulnerability Index – socioeconomic status, household composition and disability, minority status and language, and housing type and transportation – for which a score [0,1] is assigned with 0 indicating low vulnerability on that theme. The p-values are comparing the vaccinated and unvaccinated matched pregnant populations and are calculated using a Fisher’s Exact Test for categorical variables and Mann-U Whitney Test for continuous variables.

|  | No. (%) |  |  |
| --- | --- | --- | --- |
|  | Vaccinated<br>n = 26,792 | Unvaccinated<br>n =48,492 | p-value |
| **Demographics** |  |  |  |
| Race |  |  | p<0.001 |
| American Indian or Alaska Native | 205 (0.8%) | 635 (1.3%) |  |
| Asian | 4,151 (15.5%) | 3,116 (6.4%) |  |
| Black | 856 (3.2%) | 2,651 (5.5%) |  |
| Multiracial | 415 (1.5%) | 672 (7.9%) |  |
| Native Hawaiian or Pacific Islander | 212 (0.8%) | 624 (1.3%) |  |
| Other | 4,003 (14.9%) | 9,149 (18.9%) |  |
| White | 15,886 (59.3%) | 29,300 (60.4%) |  |
| Unknown | 1,064 (4.0%) | 2,345 (4.8%) |  |
| Ethnicity |  |  | p<0.001 |
| Hispanic or Latino | 6,200 (23.1%) | 13,804 (28.5%) |  |
| Not Hispanic or Latino | 20,562 (76.7%) | 34,547 (71.2%) |  |
| Unknown | 30 (0.1%) | 141 (0.3%) |  |
| Maternal Age (Years) |  |  | p<0.001 |
| 18-25 | 2,074 (7.7%) | 9,713 (20.0%) |  |
| 25-30 | 5,350 (20.0%) | 13,900 (28.7%) |  |
| 30-35 | 10,493 (39.2%) | 14,903 (30.7%) |  |
| 35-40 | 7,334 (27.3%) | 8,165 (16.8%) |  |
| 40-45 | 1,541 (5.8%) | 1,811 (3.7%) |  |
| Pregravid BMI ( kg/m2) |  |  | p<0.001 |
| Underweight | 428 (1.6%) | 704 (2.7%) |  |
| Normal | 6,725 (25.1%) | 8,495 (17.5%) |  |
| Overweight | 4,690 (17.5%) | 6,748 (13.9%) |  |
| Obese | 2,607 (9.7%) | 4,520 (9.3%) |  |
| Severely Obese | 1,273 (4.8%) | 2,314 (4.8%) |  |
| Unknown | 11,069 (41.3%) | 25,711 (53.0%) |  |
| Commercial Insurance | 18,297 (68.3%) | 19,767 (40.7%) | p<0.0001 |
| Smoker | 1,204 (4.4%) | 4,832(10.0%) | p<0.0001 |
| Illicit Drug User | 1,890 (7.1%) | 5,577 (11.5%) | p<0.0001 |
| Preterm History | 958 (3.6%) | 2,182 (4.5%) | p<0.0001 |
| Parity |  |  | p<0.001 |
| Nulliparity | 15,763 (58.8%) | 26,140 (53.9%) |  |
| Low Multiparity | 10,658 (39.9%) | 20,908 (43.1%) |  |
| Grand Multipara | 371 (1.4%) | 1,444 (3.0%) |  |
| Gravidity |  |  | p<0.001 |
| Nulligravidity | 8,811 (32.9%) | 14,541 (30.0%) |  |
| Low Multigravidity | 16,954 (63.3%) | 30,629 (63.2%) |  |
| Grand Multigravidity | 1,027 (3.8%) | 3,322 (6.9%) |  |
| **Comorbidities** |  |  |  |
| Chronic Diabetes | 2,154 (8.0%) | 3,094 (6.4%) | p<0.0001 |
| Chronic Hypertension | 525 (2.0 %) | 738 (1.5%) | p<0.0001 |
| Gestational Diabetes | 1,983 (7.4%) | 2,826 (5.8%) | p<0.0001 |
| Gestational Hypertension | 1,161(4.3%) | 2,017 (4.2%) | p=0.26 |
| Preeclampsia | 774 (2.9%) | 1,342 (2.8%) | p=0.33 |
| Severe Preeclampsia | 28 (0.1%) | 37 (0.1%) | p=0.24 |
| **Birth Characteristics** |  |  |  |
| Fetal Sex |  |  | p<0.001 |
| Female | 13,696 (51.1%) | 24,929 (51.4%) |  |
| Male | 13,046 (48.7%) | 23,388 (48.2%) |  |
| Unknown | 50 (0.2%) | 175 (0.4%) |  |
| Mode of Delivery |  |  | p<0.0001 |
| C-Section | 8,510 (31.8%) | 14,343 (30.0%) |  |
| Vaginal | 18,231 (68.0%) | 34,034 (70.2%) |  |
| Unknown | 51 (0.2%) | 115 (0.2%) |  |
| **Geographical Features** |  |  |  |
| Rural/Urban Categorization |  |  | p<0.001 |
| Rural | 204 (0.8%) | 804 (1.7%) |  |
| Small Town | 205 (0.8%) | 1,020 (2.1%) |  |
| Micropolitan | 763 (2.8%) | 2,237 (4.6%) |  |
| Metropolitan | 21,940 (81.9%) | 37,095 (76.5%) |  |
| Missing | 3,680 (13.7%) | 7,336 (15.1%) |  |
| Socioeconomic Status |  |  | p<0.001 |
| 1st Quintile (0.0 - 19.9%) | 6,784 (25.3%) | 14,574 (30.1%) |  |
| 2nd Quintile (20.0 - 39.9%) | 6,166 (23.0%) | 9,731 (20.1%) |  |
| 3rd Quintile (40.0 - 59.9%) | 4,804 (17.9%) | 10,348 (21.3%) |  |
| 4th Quintile (60.0 - 79.9%) | 3,564 (13.3%) | 8,628 (17.8%) |  |
| 5th Quintile (80.0% - 100.0%) | 1,779 (6.6%) | 5,213 (10.8%) |  |
| Missing | 3,695 (13.8%) | 7,377 (15.2%) |  |
| Household Composition and Disability |  |  | p<0.001 |
| 1st Quintile (0.0 - 19.9%) | 9,172 (34.2%) | 18,155 (37.4%) |  |
| 2nd Quintile (20.0 - 39.9%) | 5,644 (21.1%) | 9,628 (19.9%) |  |
| 3rd Quintile (40.0 - 59.9%) | 4,004 (14.9%) | 8,143 (16.8%) |  |
| 4th Quintile (60.0 - 79.9%) | 2,468 (9.2%) | 6,701 (13.8%) |  |
| 5th Quintile (80.0% - 100.0%) | 1,832 (6.8%) | 5,867 (12.1%) |  |
| Missing | 3,672 (13.7%) | 7,318 (15.1%) |  |
| Minority Status and Language |  |  | p<0.001 |
| 1st Quintile (0.0 - 19.9%) | 1,430 (5.3%) | 10,179 (21.0%) |  |
| 2nd Quintile (20.0 - 39.9%) | 3,389 (12.6%) | 6,274 (12.9%) |  |
| 3rd Quintile (40.0 - 59.9%) | 4,997 (18.7%) | 8,416 (17.4%) |  |
| 4th Quintile (60.0 - 79.9%) | 7,968 (29.7%) | 13,045 (26.9%) |  |
| 5th Quintile (80.0% - 100.0%) | 5,336 (19.9%) | 10,580 (21.8%) |  |
| Missing | 3,672 (13.7%) | 7,318 (15.1%) |  |
| Housing Type and Transportation |  |  | p<0.001 |
| 1st Quintile (0.0 - 19.9%) | 2,729 (10.2%) | 11,397 (23.5%) |  |
| 2nd Quintile (20.0 - 39.9%) | 4,698 (17.5%) | 8,506 (17.5%) |  |
| 3rd Quintile (40.0 - 59.9%) | 4,353 (16.2%) | 7,086 (14.6%) |  |
| 4th Quintile (60.0 - 79.9%) | 4,312 (16.1%) | 8,291 (17.1%) |  |
| 5th Quintile (80.0% - 100.0%) | 7,005 (26.1%) | 13,214 (27.2%) |  |
| Missing | 3,700 (13.8%) | 7,387 (15.2%) |  |

Supervised learning models were trained using 20 demographic, comorbidity, and geographical features to predict vaccination status at delivery (Supplemental Table 4). Model performance was evaluated using a held-out test dataset (Supplemental Table 6). The highest performing model was the gradient boosting regression model, which had an area under the curve (AUC) of 0.80 and R^2^ of 0.25 (Figure 1C; Supplemental Table 6). The six most important features for the model were commercial insurance status, community housing composition and disability vulnerability level, age, community socioeconomic vulnerability level, pregravid BMI, and rural/urban categorization (Supplemental Figure 2-3). These six features alone were used to train a limited gradient boosting regression model that performed similarly to the original model with an AUC of 0.79 and R^2^ of 0.25 (Figure 1C-D; Supplemental Table 6).

These 20 features were used to create an unvaccinated matched cohort using propensity score matching of k-nearest neighbors with replacement (k=1; Supplemental Table 4). This resulted in an unvaccinated matched cohort that more closely resembled the vaccinated cohort in composition (Supplemental Table 7). There was a reduction in the absolute standard mean difference between the vaccinated and unvaccinated matched cohorts for all the covariates except minority status and language vulnerability and rural/urban categorization (Supplemental Figure 4). After matching the absolute standard mean difference between vaccinated and unvaccinated matched cohorts was less than 0.09 for all 20 features.

### Evaluating the maternal COVID-19 infection rate in vaccinated people

The number of pregnant people reaching full vaccination status peaked in May of 2021 (Figure 2A). A drop-off of initiation of the COVID-19 vaccination series was observed during the first trimester, but there was a higher rate of COVID-19 vaccination in the second and third trimester (Figure 2B). Over a five-month period of pregnancy, vaccinated people (n=6,119) had lower rates of COVID-19 infection compared to unvaccinated people matched on conception date (n=6,119; p<0.01), which controls for gestational timing and pandemic timing. (Figure 2C).

**Figure 2.**
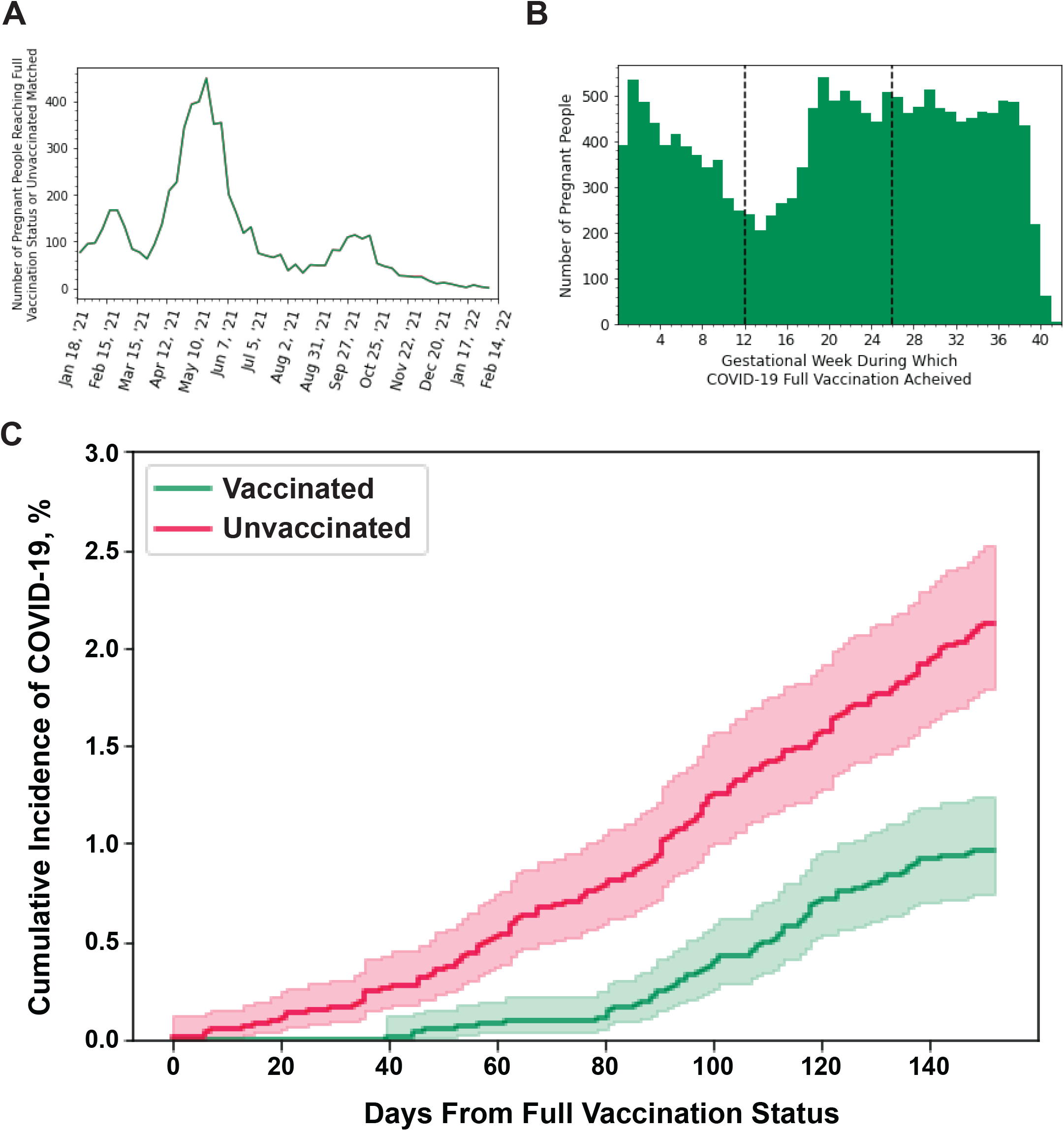
Effectiveness of mRNA vaccine at preventing COVID-19 infections in pregnant people. **A:** Weekly count of the number of people reaching full vaccination status (two weeks following second mRNA dose in the initial vaccination series) during pregnancy **B:** Histogram of the gestational week people achieved full vaccination status. **C:** Percentage of people plus the 95% CI that had COVID-19 infections over five months (152 days) of pregnancy in mRNA vaccinated (green) or unvaccinated (red). T0 is at the day of full vaccination (2 weeks after the last dose of the initial vaccination series). Unvaccinated people were matched to vaccinated people on conception date and an unvaccinated person T0 is the full vaccination date of the matched vaccinated person. The p-values was calculated by a log-rank test. Vaccinated n=6,119; Unvaccinated: n=6,119. Vaccinated vs Unvaccinated: p<0.01.

Most maternal COVID-19 cases were in unvaccinated people; few vaccinated or boosted individuals had breakthrough infections prior to Omicron obtaining dominance (Figure 3A; Supplemental Table 8). There were 916 vaccinated people (191 of whom were boosted at the time of infection), 3,394 unvaccinated people, and 1,178 unvaccinated matched people who had maternal COVID-19 infections. A significantly higher proportion of COVID–19 infections occurred in the vaccinated cohort during Omicron dominance (p<0.001; Figure 3A-B). There were fewer maternal COVID-19 cases in boosted people even during Omicron dominance (Figure 3A). The proportion of people infected with COVID-19 during pregnancy is significantly higher in unvaccinated people than either vaccinated (p<0.0001) or boosted (p<0.0001) people. The proportion of people with a positive COVID-19 test during pregnancy was significantly lower in those who had received a booster than those that are vaccinated, but not boosted (p<0.0001). Finally, the rate of maternal COVID-19 infection was significantly lower in the vaccinated cohort (0.034, 95% CI=[0.032, 0.036]) compared to both the unvaccinated matched cohort (0.044, 95% CI=[0.042, 0.047]; p<0.0001) and unvaccinated cohort (0.07, 95% CI=[0.068, 0.072]; p<0.0001; Figure 3E, far left panel).

**Figure 3.**
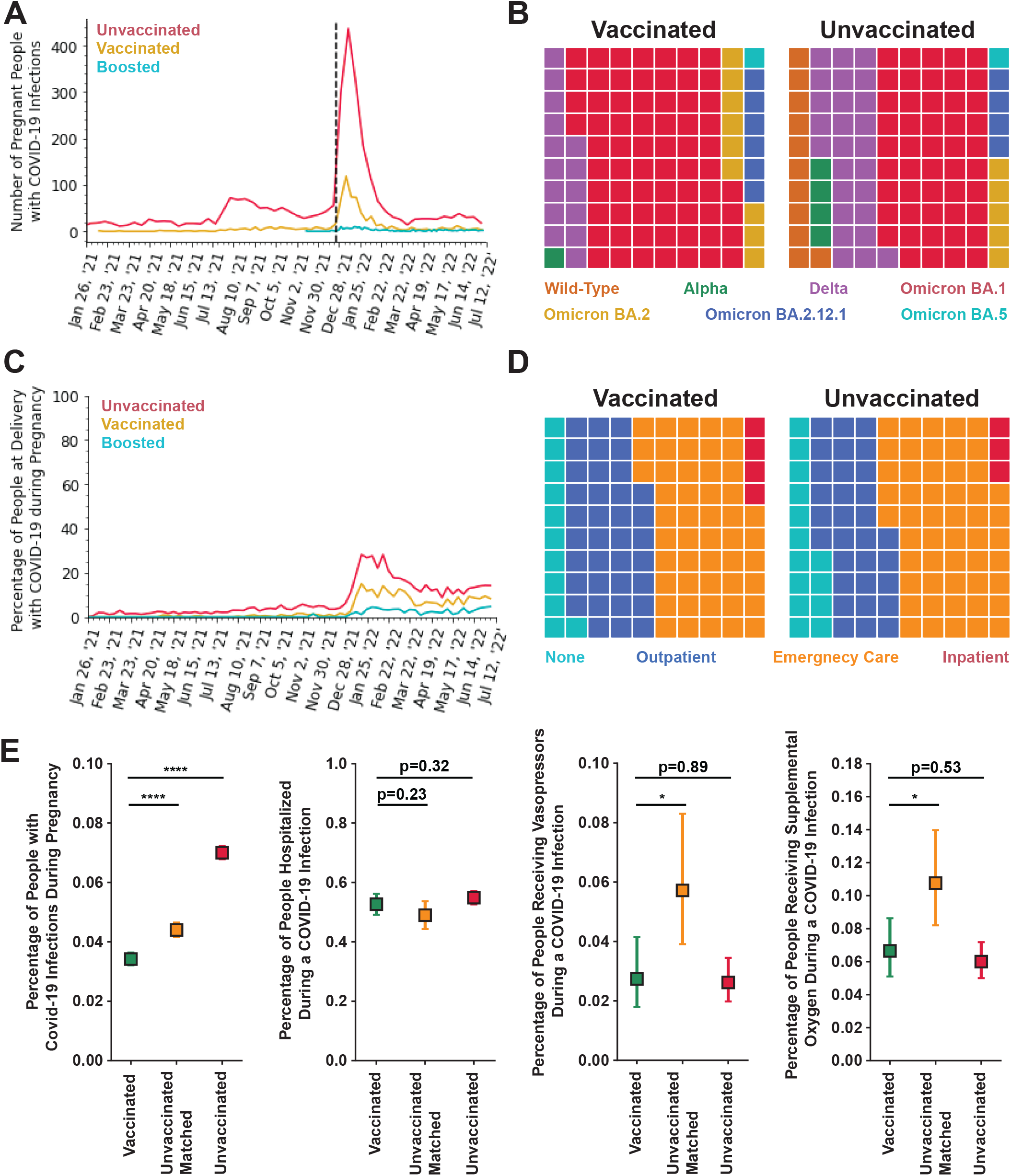
Evaluation of maternal COVID-19 infections by vaccination status. **A:** Weekly counts of COVID-19 infection from January 26, 2021 through July 11, 2022 by vaccination status - unvaccinated (red; n=3,119), vaccinated, two mRNA doses only (gold; n=723), or boosted (teal; n=193). The dotted line indicates the day when Omicron BA.1 became the dominant variant in the western U.S. (December 25, 2021). **B:** Waffle plot of the dominant variant – Wild-Type (orange), Alpha (green), Delta (purple), Omicron BA.1 (red), Omicron BA.2 (gold), Omicron BA.2.12.1 (blue), or Omicron BA.5 (teal) – at time of maternal COVID-19 infection for Vaccinated (left panel; n=916) or Unvaccinated (right panel; n=3,119). p<0.001; Fishers Exact Test. **C:** Percentage of people at delivery that had a COVID-19 infection during their pregnancy from January 26, 2021 through July 11, 2022 by vaccination status - unvaccinated (red; n=48,492), vaccinated, two mRNA doses only (gold; n=19,176), or boosted (teal; n=7,616) - for Vaccinated (left; n=916) and Unvaccinated (right; n=3,119). The p-values were calculated by ANOVA. Unvaccinated vs Vaccinated, two mRNA doses only: p<0.0001; Unvaccinated vs Boosted: p<0.0001; Vaccinated, 2 mRNA doses only vs Boosted: p<0.0001. **D:** Waffle plot of the type of care sought during a maternal COVID-19 infection after omicron achieved dominance (infections after December 25, 2021) – None (teal), Outpatient (blue), Emergency Care (orange), or Inpatient (red) – for Vaccinated (left panel; n=767) or Unvaccinated (right panel; n=1,833) individuals. p<0.05; Fishers Exact Test. **E:** Percentage of people with 95% CI for vaccinated (green), unvaccinated matched (yellow), and unvaccinated (red) that have had a maternal COVID-19 infection (far left panel; vaccinated n=916; unvaccinated matched n=1178; unvaccinated n=3,119) or had the following outcomes during an active maternal COVID-19 infection after Omicron achieved dominance (infection after December 24, 2021; vaccinated n=767; unvaccinated matched n=437; unvaccinated n=1,833): hospitalization (center left panel), vasopressor use (center right panel) or supplemental oxygen use (far right panel). The 95% CI was calculated by Wilson Score Interval. p-value was calculated by Mann-U Whitney Test for continuous variables or Fisher’s Exact Test for categorical variables. * p<0.05; **** p<0.0001.

### Evaluating maternal COVID-19 infection severity in vaccinated people

Vaccinated people with a maternal COVID-19 infection during Omicron dominance were less likely to require emergency care and more likely to receive outpatient care than people in the unvaccinated cohort (p<0.05; Figure 3D). Vaccinated people also had a significantly different distribution of type of care for a maternal COVID-19 infection than unvaccinated matched pregnant people, with more vaccinated people seeking care (p<0.001). Surprisingly, the overall rate of hospitalization rate (emergency care or inpatient) for maternal COVID-19 infections was the same in vaccinated (0.527, 95% CI=[0.491, 0.562]) compared to both unvaccinated matched (0.490, 95% CI=[0.443, 0.537], p=0.23), and unvaccinated (0.549, 95% CI=[0.526, 0.572], p=0.32) cohorts (Figure 3E, center left panel). There were three deaths during maternal COVID-19 infections in the unvaccinated cohort (0.1%; 3/3,394) and three in the unvaccinated matched cohort (0.3%.; 3/1,178). No maternal deaths were observed in any other cohorts. Two of these deaths occurred during Delta dominance and the third occurred following an infection contracted shortly before Delta became the predominant variant.

Vaccinated (0.027, 95% CI=[0.018, 0.042]) pregnant people were significantly less likely to require vasopressors during a maternal COVID-19 infection than unvaccinated matched (0.057, 95% CI=[0.039, 0.083], p<0.05), but not unvaccinated people (0.026, 95% CI=[0.020, 0.035], p=0.89; Figure 3E, center right panel). In addition, vaccinated people (0.067, 95% CI=[0.051, 0.086]) were significantly less likely to require supplemental oxygen during a maternal COVID-19 infection than those in the unvaccinated matched cohort (0.108, 95% CI=[0.082, 0.140], p<0.05), but not the unmatched unvaccinated cohort (0.060, 95% CI=[0.050, 0.072], p=0.53; Figure 3E, far right panel). There was no difference in the COVID-19 severity of infection between vaccinated and either unvaccinated matched or unvaccinated people (Supplemental Table 9).

### Comparing the efficacy of three types of vaccine

Evaluation of the maternal COVID-19 infection and related hospitalization for people with one mRNA vaccine dose, Ad26.COV2.S Johnson & Johnson vaccine, or COVID-19-induced immunity compared to unvaccinated people are reported (Supplemental Results; Supplementary Figure 5). Evaluation of the efficacy of mRNA-1273 Moderna compared to BNT162b2 Pfizer-BioNTech vaccines is also reported (Supplemental Results; Supplementary Figure 6).

### Evaluating birth outcomes in vaccinated people

Vaccinated (0.003, 95% CI=[0.003, 0.004]) people had significantly lower rates of stillbirth than both unvaccinated (0.005, 95% CI=[0.005, 0.006] p<0.001) and unvaccinated matched (0.005, 95% CI=[0.004, 0.006], p<0.01) people (Figure 4A). Vaccinated people also had significantly lower rates of PTB (0.081, 95% CI=[0.078, 0.084]) and VLBW (0.010, 95% CI=[0.009, 0.012]) than unvaccinated (PTB: 0.086, 95% CI=[0.084, 0.089], p<0.05; VLBW: 0.013, 95% CI=[0.012, 0.014, p<0.01]) but not unvaccinated matched people (PTB: 0.079, 95% CI=[0.076, 0.082], p=0.35; VLBW: 0.011, 95% CI=[0.010, 0.012, p=0.31]; Figure 4A). There was no difference in the SGA rate in vaccinated (0.133, 95% CI=[0.129, 0.137]) people compared to either unvaccinated (0.134, 95% CI=[0.131, 0.137], p=0.67) or unvaccinated matched (0.133, 95% CI=[0.130, 0.138], p=0.79) people (Figure 4A). In the vaccinated cohort there was a significant decrease in the proportion of babies born prematurely that were born very or extremely preterm (<32 weeks gestation) compared to unvaccinated matched (p<0.05) and unvaccinated (p<0.001) people (Figure 4B). Babies born to vaccinated people have significantly higher gestational age at delivery than unvaccinated people, but comparable gestational age as unvaccinated matched people (Supplemental Table 11).

**Figure 4.**
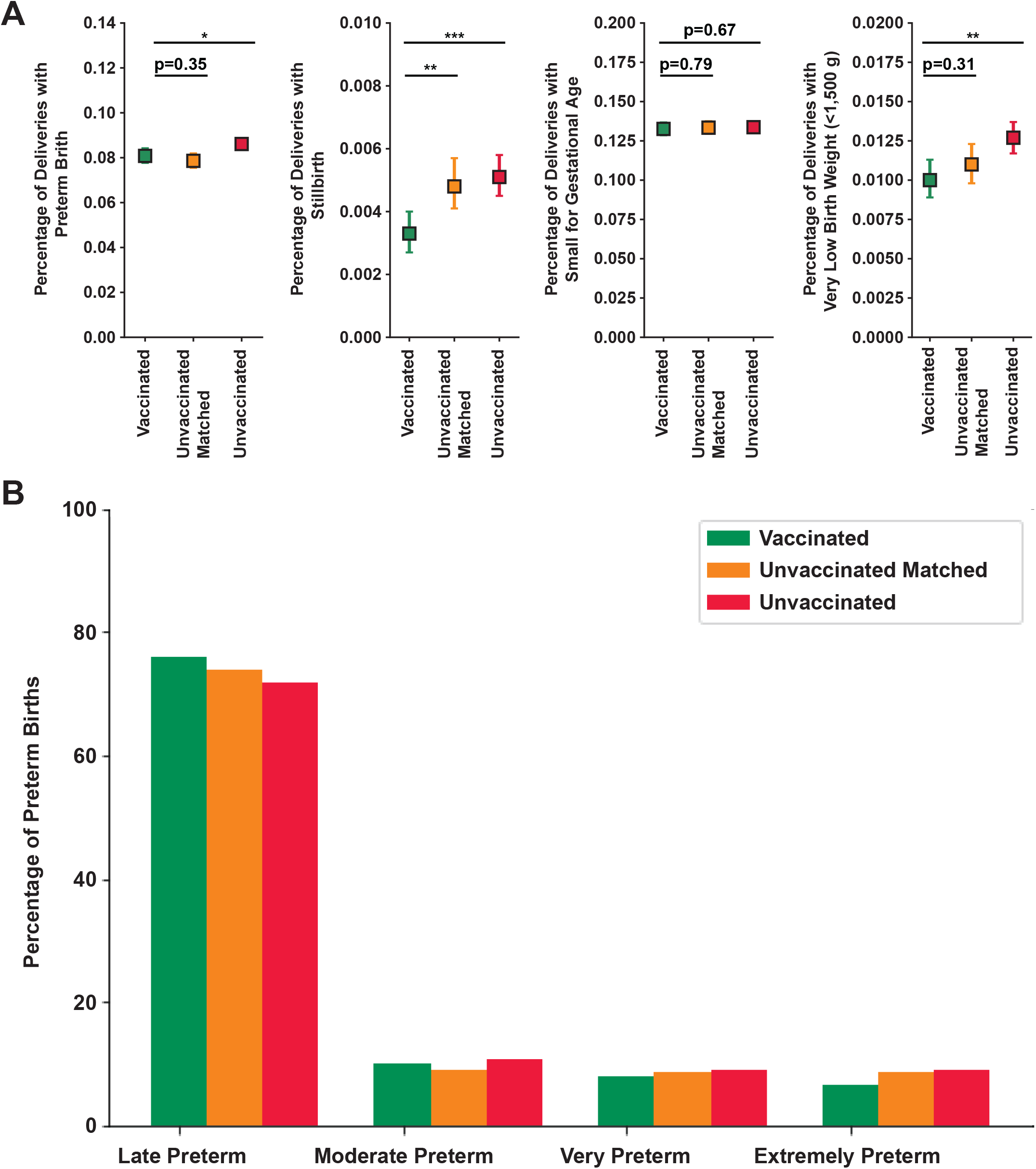
Birth outcomes by vaccine status. **A:** Percentage of people with 95% CI for vaccinated (green; n=26,792), unvaccinated matched, (yellow n=26,790) or unvaccinated (red; n=48,492) that had a preterm delivery (far left panel), stillbirth (center left panel), small for gestational age (bottom 10th fetal growth percentile at delivery; center right panel) or very low birth weight (<1,500 g; far right panel). The 95% CI was calculated by Wilson Score Interval and the p-values by Mann-U Whitney Test. * p<0.05; ** p <0.01; *** p<0.001. **B:** Proportion of preterm deliveries that were late preterm (<37 and >34 weeks gestation), moderate preterm (<34 and >32 weeks gestation), very preterm (<32 and >28 weeks gestation) or extremely preterm (<28 weeks gestation) for vaccinated (green; n=2,166), unvaccinated matched (yellow; n=2,106), and unvaccinated (red; n=4,176) people. P-values were calculated by Fisher’s Exact Test. Vaccinated vs Unvaccinated Matched: p<0.05; Vaccinated vs Unvaccinated: p<0.001.

### Evaluating booster efficacy compared to two mRNA COVID-19 doses in pregnant people

Most people that received a third mRNA COVID-19 dose received it substantially after the CDC recommended time of five months following the second dose (Figure 5A). People that were boosted had significantly lower rates of maternal COVID-19 infection (0.025, 95% CI=[0.022, 0.029]) than those who were vaccinated, but not boosted (0.050, 95% CI=[0.048, 0.57], p<0.0001; Figure 5B, left panel). The timing from full vaccination status to breakthrough infection was also significantly longer in boosted people than in vaccinated, but not boosted people (p<0.0001; Figure 5C). Boosted people (0.449, 95% CI=[0.38, 0.521]) were less likely to be hospitalized with a maternal COVID-19 infection during Omicron dominance than vaccinated, but not boosted people (0.548, 95% CI=[0.48, 0.57], p<0.05; Figure 5B, right panel). Boosted people also had a difference in the distribution of the number of outpatient and total medications, but not the number of inpatient medications administered during an active covid infection compared to vaccinated but not boosted people (Supplemental Table 12).

**Figure 5.**
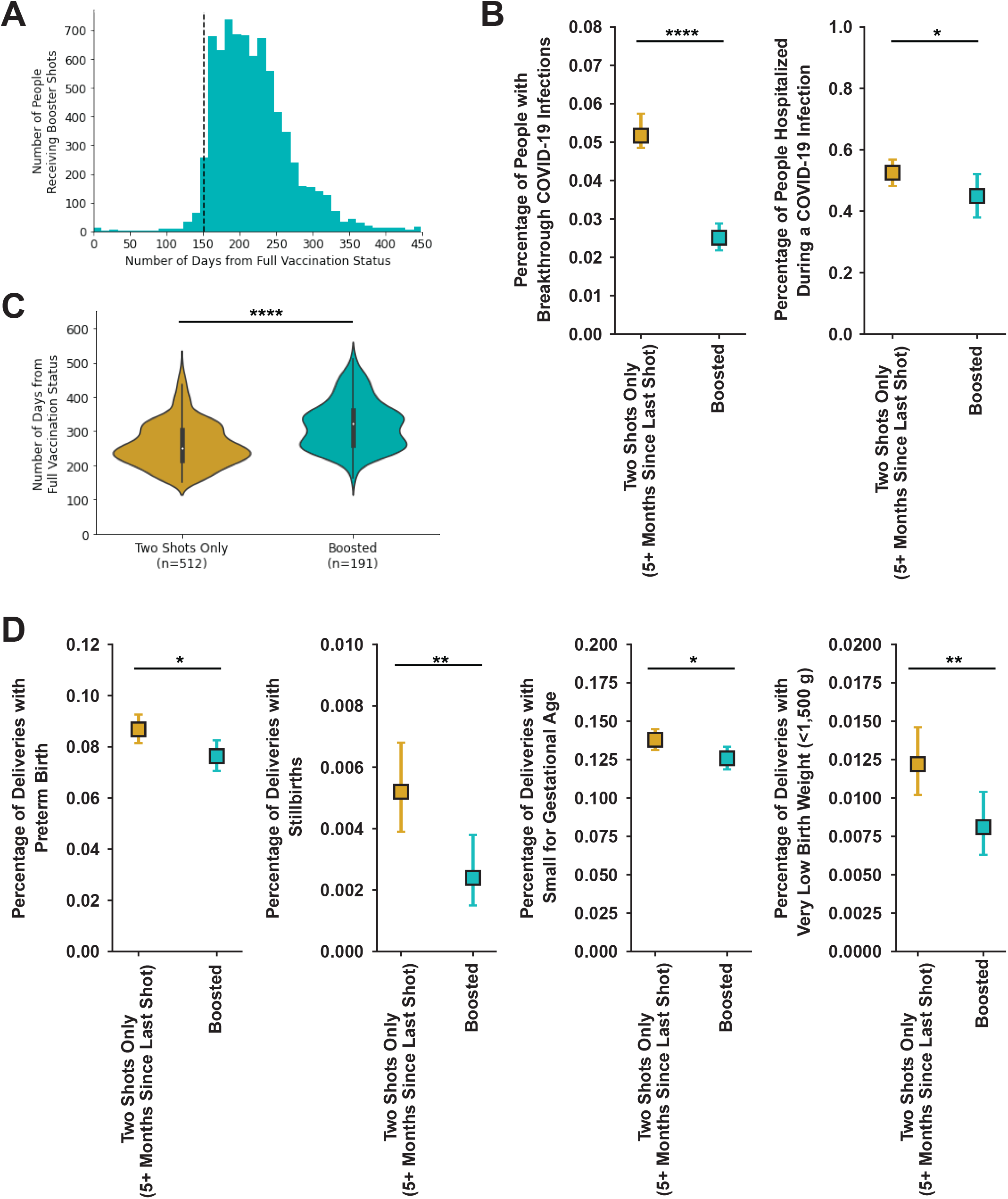
Evaluation of maternal-fetal outcomes by booster status. **A:** Histogram of number of days between last COVID-19 vaccine dose and booster dose. The black line at day 152 indicates (five months) and is the recommended timing of the third COVID-19 dose (booster) by the CDC. n=7,616. **B:** Violin plot of the time from day of achieving full vaccination status to maternal breakthrough COVID-19 infection for vaccinated people with no booster dose greater than five months after last COVID-19 vaccine dose (gold; n=512) or individuals that received a third mRNA dose at least five months after completing the initial series (teal; n=191). **C:** Percentage of people with 95% CI for vaccinated but not boosted (gold) and boosted (teal) with maternal COVID-19 infection (left panel; two mRNA dose n=9,708; boosted n=7,558) or were hospitalized during a maternal COVID-19 infection after Omicron became dominant (infection after December 24, 2021; two mRNA doses n=495; boosted n=187). **D:** Percentage of people with 95% CI for vaccinated but not boosted (gold; n=9,708) and boosted teal; n=7,558) that had a preterm delivery (far left panel), stillbirth (center left panel), small for gestational age (bottom 10th fetal growth percentile at delivery; center right panel) or very low birth weight (<1,500 g; far right panel). The 95% CI calculated by Wilson Score Interval and p-value by either Mann-U Whitney Test for continuous variables or Fisher’s Exact Test for categorical variables. * p<0.05; ** p<0.01; **** p<0.0001.

### Evaluating birth outcomes between boosted and vaccinated (but not boosted) people

Boosted people had significantly lower rates of PTB (0.076, 95% CI=[0.071, 0.083]), stillbirth (0.002, 95% CI=[0.002, 0.004]), SGA (0.126, 95% CI=[0.119, 0.133]), and VLBW (0.008, 95% CI=[0.006, 0.010]) compared to those that are vaccinated, but not boosted (PTB: 0.089, 95% CI=[0.081, 0.093], p<0.05; stillbirth: 0.005, 95% CI=[0.004, 0.007], p<0.01; SGA: 0.138, 95% CI=[0.131, 0.145], p<0.05; VLBW: 0.012, 95% CI=[0.010, 0.015], p<0.01; Figure 5D). Babies born to boosted people are significantly less likely to have a LBW and have significantly higher birth weight than those born to vaccinated, but not boosted people (Supplemental Table 13).

## Discussion

We examined the impact of COVID-19 vaccination on maternal-fetal outcomes and found the following results. Compared to unvaccinated people, vaccinated people were significantly more likely to be older, have a higher BMI, have commercial insurance, live in an urban area, and live in areas with lower vulnerability index values for socioeconomics, and housing composition and disability. Vaccinated people had a lower rate of maternal COVID-19 infection and this difference remained even after controlling for 20 covariates using propensity score matching. All three maternal deaths associated with COVID-19 infection were in unvaccinated individuals. Breakthrough cases in vaccinated individuals occurred disproportionately during the time of Omicron dominance. Vaccinated people compared to unvaccinated matched people had similar rates of hospitalization, but significantly lower rates of supplemental oxygen use and vasopressor use during a maternal COVID-19 infection during Omicron dominance. Vaccinated people had better or comparable birth outcomes than unvaccinated people in a matched cohort: lower rates of stillbirth and similar rates of PTB, SGA, and VLBW. Furthermore, receiving a third mRNA COVID-19 booster dose at least five months following completion of the two-dose mRNA vaccination series was associated with significantly lower rates of maternal COVID-19 infection, COVID-19 related hospitalization, PTB, stillbirth, SGA, and VLBW compared to individuals that only received two mRNA doses. Altogether, this supports the conclusion that COVID-19 mRNA vaccination offers protection against adverse maternal-fetal outcomes, and that third boosters support statistically and clinically significant improvement in maternal-fetal outcomes.

The results of COVID-19 vaccination and infection are comparable to those previously reported in non-pregnant adults. Both one-dose and two-dose mRNA vaccination protects against infection, with two doses conferring additional protection.^33^ Also, rates of COVID-19 breakthrough infections were lower in people that received three-doses compared to two-doses of mRNA vaccine.^34–37^. Finally, vaccine efficacy has also reported to be lower against infection by the Omicron variant compared to the Delta variant.^34,35,37,38^ During rapidly evolving pandemic circumstances, research in the pregnant population lags research performed in non-pregnant adults. However, the consistency between the results of COVID-19 vaccination in pregnant people and what has been previously reported in the non-pregnant adult population gives confidence to using information obtained in the non-pregnant adult population to guide recommendations for the pregnant population when studies specific to the pregnant population are unavailable.

We report neutral or better birth outcomes in people that were vaccinated at delivery compared to unvaccinated people even after controlling for 20 demographic, lifestyle, clinical, and geographical characteristics. Consistent with previous studies, we report no difference in the rate of PTB or SGA between vaccinated and unvaccinated individuals.^25–27^ We also report a significant decrease in the proportion of PTBs that were very or extremely preterm (<32 weeks gestation) in the vaccinated population which is similar to the decrease in babies born to vaccinated people prior to 32 weeks of gestation observed in Sweden, but not Norway.^27^ Unlike previous studies, we observed a significantly lower rate of stillbirth in vaccinated individuals.^9,27^ Previous studies deployed descriptive statistics (prevalence rate or odds ratio with the 95% CI), whereas we conducted additional quantitative analyses to evaluate these outcomes. Our study also had a longer observation period and included patients delivering during and after the Omicron BA.1, BA.2, and BA.2.12.1 waves, and during the beginning of the Omicron BA.5 wave. Maternal COVID-19 infection is known to increase the risk for PTB, stillbirth, SGA and decreased birth weight.^5–10^ COVID-19 vaccination reduces the risk of maternal COVID-19 infection, and this decreased infection rate in vaccinated may explain the decrease rates in these negative birth outcomes.^18–20^ These protective effects might be explained in part by healthy vaccine bias.

For the first time, we report maternal-fetal outcomes of the third booster mRNA COVID-19 dose in pregnant people. We observed significantly decreased rates of COVID-19 infection, PTB, stillbirth, SGA, LBW, and VLBW in vaccinated people that received a booster dose. This provides strong support for pregnant people to receive a third booster dose. Vaccine efficacy has been shown to wane as timing from administration of the second or third dose of the mRNA vaccine increases,^35–37^ and in older adults (>60 years old), rates of infection and severe illness were lower in people that received four versus three doses.^39^ This supports that receiving additional vaccination doses increases vaccine effectiveness. Therefore, future studies should explore the benefit of a fourth dose of COVID-19 vaccine in pregnant people.

A strength of this study is that this is a multicenter study using detailed medical records from an integrated healthcare system that serves a large population across diverse geographic regions, enabling the most in-depth and complete results to date on the impact of COVID-19 vaccination and booster doses on maternal-fetal outcomes. Our results on positive maternal-fetal outcomes replicate even after controlling for 20 demographic, lifestyle, clinical, and geographical characteristics between the vaccinated and unvaccinated populations. Both mRNA-1273 Moderna and BNT162b2 Pfizer-BioNTech are associated with positive maternal-fetal outcomes. Given these results, pregnant people’s increased risk for morbidity and mortality from COVID-19 infection, and maternal COVID-19 infection associated with increased risk of negative birth outcomes, pregnant people should be encouraged to become vaccinated and stay current with their COVID-19 vaccinations as part of routine prenatal care.^3,8,9^ This evidence supports existing recommendations by health organizations around the world, and can inform policy about vaccine prioritization.

This study has some limitations. It was a retrospective cohort study without randomized control of the cohort composition, and there were significant differences in the demographics, comorbidities, social determinants of health, and geographical factors between the vaccinated and unvaccinated cohorts. We controlled for 20 factors using propensity score matching and reduced the difference for 18. However, some small level of imbalance remained. There may also be additional covariates that impact maternal-fetal outcomes for which we did not account. In addition, this study was conducted without validation at an independent healthcare system. However, the size and diversity of PSJH (51 hospitals and 1085 clinics across seven states) helps mitigate concerns of generalizability. Further, studies investigating the impact of COVID-19 vaccination on maternal-fetal outcomes necessitates simultaneous consideration of three distinct timelines: gestational age, pandemic timing and length of time from last vaccine dose. Both vaccine effectiveness and disease severity have been shown to be variant dependent,^34,35,37,38^ with Delta being associated with more severe disease than Alpha or Omicron^38^ in both vaccinated and unvaccinated populations. We account for this by asking questions of note, such as the COVID-19 infection rate in vaccinated versus unvaccinated people, in multiple ways to partially account for each or all of these temporal factors alongside other confounding variables. In other cases, such as the impact of COVID-19 vaccination status on maternal COVID-19 disease severity, we limit our scope to Omicron COVID-19 infections only. We did not have genotype results for infections, so instead limited cases to those occurring Omicron became dominant (>50% of infections) in the western USA based on genotype surveillance.^17^ In this way, we attempt to supply a more complete picture of the impact of COVID-19 vaccination on maternal-fetal outcomes, despite the complexity of the issue.

This multicenter retrospective cohort study found that vaccinated pregnant people were older, had higher pregravid BMI, were more likely to have commercial insurance, and were more likely to live in urban areas and areas with lower social vulnerability than unvaccinated people. Vaccinated people had a decreased rate of a maternal COVID-19 infection than unvaccinated people. Unexpectedly, they had similar levels of COVID-19 related hospitalization, but they did have lower rates of some indicators of COVID-19 severity. Vaccinated people had decreased rates of stillbirth and comparable rates of PTB, SGA, and VLBW compared to unvaccinated people. Furthermore, boosted people had decreased rates of maternal COVID-19 infection, COVID-19 related hospitalization, PTB, stillbirth, SGA, and VLBW compared to people that were vaccinated, but not boosted. Altogether, this indicates that COVID-19 mRNA vaccination promotes positive maternal-fetal outcomes with booster doses conveying additional protection. Therefore, pregnant people should be recommended for initial COVID-19 vaccination and to stay current with their COVID-19 vaccination schedule as part of routine prenatal care.

## Data Availability

All clinical logic has been shared. Results have been aggregated and reported within this paper to the extent possible while maintaining privacy from personal health information (PHI) as required by law. All data is archived within PSJH systems in a HIPAA-secure audited compute environment to facilitate verification of study conclusions.

## Acknowledgements

This work was funded by the National Institute for Child Health & Human Development grant HD091527 as well as William O. and K. Carole Ellison Foundation. We are grateful to PSJH for sharing their data engineering expertise and computational resources. We would also like to acknowledge SNOMED International for developing and maintaining SNOMED-CT©.

## Author contributions

SNP, JJH, LH, and NDP conceptualized the study. SNP was responsible for developing the study methodology and performing data visualization. SNP, YMH, and RTR performed data cleaning and transformation. SNP did the data analyses, including statistical analysis and machine learning. SNP and YMH were involved in the investigation. SNP, JJH, LH, NDP, YMH, and TS were involved in data interpretation. JJH and LH supervised the study implementation. Administrative and material support was provided by JJH and LH. Funding was acquired by NDP, LH, and SNP. SNP prepared the original draft of the manuscript with critical revision of the manuscript for important intellectual content provided by JJH, LH, and NDP. All authors reviewed and approved of the final version of the manuscript. JJH, SNP, and YMH had full access to the data in the study. All authors had final responsibility for the decision to submit for publication.

