## Supplementary Material for "The effect of COVID-19 vaccination and booster on maternal-fetal outcomes: a retrospective multicenter cohort study"

### Contents


Supplemental Methods 2

Supplemental Results 5

Supplemental Figure 1. Flow diagram of cohort selection. 7

Supplemental Figure 2. Contribution of features towards predicting vaccination status. 8

Supplemental Figure 3. Shapley permutation explainer of the feature contribution towards classifying vaccination status. 9

Supplemental Figure 4. Propensity score matching reduces the differences in covariates between vaccinated and unvaccinated cohorts. 10

Supplemental Figure 5. Rates of maternal COVID-19 infection and subsequent hospitalization in populations with immunity. 11

Supplemental Figure 6. Effectiveness of mRNA-1273 Moderna and BNT162b2 Pfizer-BioNTech at preventing COVID-19 infections. 12

Supplemental Table 1. Vaccination status definitions. 13

Supplemental Table 2. Feature definitions for demographic, birth characteristic, and geographical features. 14

Supplemental Table 3. Pregnancy-related conditions SNOMED definitions. 15

Supplemental Table 4. Feature definitions for models predicting vaccination status at delivery. 17

Supplemental Table 5. Demographic, comorbidity, and geographical features association with vaccination status at delivery. 18

Supplemental Table 6. Performance metrics of machine learning models predicting vaccination status at delivery. 18

Supplemental Table 7. Demographic, comorbidities, birth characteristics, and geographical features of unvaccinated matched people. 19

Supplemental Table 8. Dominant variant timing in the western United States. 21

Supplemental Table 9. Additional outcomes related to maternal COVID-19 infection. 22

Supplemental Table 10. Demographic, comorbidities, birth characteristics, and geographical features of pregnant people vaccinated with Moderna vs Pfizer. 23

Supplemental Table 11. Additional birth outcomes by vaccination status. 25

Supplemental Table 12. Additional outcomes related to maternal COVID-19 infection. 25

Supplemental Table 13. Additional birth outcomes by boosted status. 26

Supplemental References 26

### Supplemental Methods

*Descriptive statistical analyses*

The composition of each cohort's demographic features, comorbidities, birth characteristics, and geographical features were represented as proportions (Table 1). Demographic and birth characteristics were defined as binary or categorical variables (Supplemental Table 2). Comorbidities were identified by patient diagnosis codes, using SNOMED-CT© and were represented as binary variables (Supplemental Table 3). To define geographical features for patients, we mapped individual patients to U.S. census tracts based on their recorded address. Patients were then mapped to CDC Social Vulnerability Index (SVI) themes and U.S. Department of Agriculture Economic Service (USDA ERS) Rural-Urban Commuting Area (RUCA) codes.[1,2] CDC Social Vulnerability Index describes the potential risk of public health emergencies caused by external stresses on human health.[1] It ranks social vulnerability for each U.S. census tract on 15 social factors, which are summarized into four themes: socioeconomic status, household composition and disability, minority status and language, and housing type and transportation. These are represented as continuous variables [0,1] with 0 indicating a low vulnerability level on that theme. USDA ERS RUCA codes are a classification system that divides census tracts into metropolitan [0,4), micropolitan [4,7), small town [7,10), or rural [10,99) areas based on the population size and commuting flow.[2] We used the secondary RUCA codes, which were generated in 2010 and last revised in 2019, to generate a categorical variable for rural/urban categorization. The differences between cohorts were evaluated by Fisher's Exact Test using R stats (version 4.1.1) for categorical variables and Mann-U Whitney Test using Python scipy (version 1.4.1) for continuous variables.

We reported the weekly counts of deliveries and COVID-19 infections by vaccination status from January 26, 2021, to July 11, 2022. We describe the weekly proportion of people delivering that had a COVID-19 infection at some point during their pregnancy by vaccination status during this period. The difference by vaccination status between the proportions of the population with a maternal COVID-19 infection during pregnancy was evaluated using a repeated measures ANOVA test using python package statsmodels (version 0.12.2). In addition, we noted the weekly counts of people achieving full vaccination status during pregnancy from January 18, 2021, to February 11,

2022. A histogram of the counts by gestational week during which people achieved full vaccination status was generated. Another histogram reported the number of days after receiving the second COVID-19 vaccination that people received the third COVID-19 vaccine booster dose.

In addition, the type of mRNA COVID-19 vaccination received by the vaccinated population and the timing the COVID-19 vaccine doses received in relation to the person's pregnancy were reported. The COVID–19 infections the proportions that occurred during the different dominant strains and the highest level of care (none, outpatient, emergency care, or inpatient) was reported for vaccinated and unvaccinated individuals. A Fisher's exact test was performed to evaluate the difference in population distribution between these two cohorts using R stats (version 4.1.1). All graphs were generated using python package matplotlib (version 3.4.2).

*Classification models*

To evaluate which demographic or social determinants of health features were most predictive of vaccination status at delivery (vaccinated versus unvaccinated) we trained multiple supervised learning models on 20 features. These features included demographics, insurance, habits, comorbidities, and geographical features (Supplemental Table 4). Race was coded as a binary variable (not identifying = 0; identifying = 1) for the four races for which there were at least 1,000 people identifying as that race. Pregravid BMI was separated into five standard categories with missing values encoded as -1. The correlation of each feature with vaccination status was evaluated using Pearson correlation calculated by the python library scipy (version 1.6.2).

Models were generated using python package sklearn (version 1.0.2) with default settings for logistic regression, gradient boosting regression, and random forest. The models were trained on 80% of the data with 20% of the data withheld for evaluation of the final performance of the model. Model performance was evaluated using mean absolute error, mean squared error, root mean squared error, area under the curve (AUC), and $R^2$. Gini feature importance was used to assess the marginal contribution and influence of each feature on the final model providing interpretation of the machine learning models. In addition, on 1,000 randomly selected patients from the test set the Shapley additive explanations (SHAP) was used to evaluate the average marginal contribution of a feature value across all permutations of features providing insight into the degree of influence of the feature on an individual's predicted vaccination status at delivery for the gradient boosting models. A limited version of the top performing model (gradient boosting regression) was also trained using the top six most important features from the full model. Predictive models were reported following the TRIPOD guidelines.[3]

*Reverse kaplan-meier curve*

A Kaplan-Meier curve reporting the event of a COVID-19 infection was reported over a five-month (152 day) period using python package lifelines (version 0.27.0). This was reported starting at the day of full-vaccination status (14 days after the second dose) for either the mRNA-1273 Moderna or BNT162b2 Pfizer-BioNTech vaccine. Unvaccinated people were matched to vaccinated people by conception date and T0 was the date the corresponding vaccinated person reached full vaccination status. All people were pregnant for the entire five-month period. A log-rank test was used to evaluate the difference in event occurrence between these cohorts. The graph was plotted to show the percentage of people that had a COVID-19 infection starting at 0% at T0.

*Quantitative statistical analyses*

Violin plots were used to report the number of days from full vaccination status to breakthrough status for people vaccinated with mRNA-1273 Moderna or BNT162b2 Pfizer-BioNTech as well as those that were boosted or vaccinated, but not boosted. Differences in these populations were evaluated using a Mann-U Whitney test using python package scipy (version 1.6.2). Rates in COVID-19 infections and related hospitalization along with the 95% Confidence Interval (CI) were calculated using the Wilson Score Interval using python package statsmodels (version 0.12.2) for unvaccinated, vaccinated, one mRNA dose, vaccinated with Ad26.COV2.S Johnson & Johnson, and COVID-19-induced immunity cohorts. Rates of preterm birth (born <37 weeks gestation), stillbirth (fetal demise at ≥20 weeks gestation), low birth weight (birth weight <2,500 g), very low birth weight (birth weight <1,500 g), and small for gestational age (bottom 10th fetal growth percentile at delivery) were reported for unvaccinated, vaccinated, and boosted cohorts. The timing of premature birth was considered by dividing it into four categories. Term birth is defined as birth >37 weeks gestation, late preterm birth as <37 and ≥34 weeks gestation, moderate preterm birth as <34 and ≥32 weeks gestation, very preterm birth as <32 and ≥28 weeks gestation, and extremely preterm birth <28 weeks gestation. Fetal growth percentile was calculated using the World Health Organization (WHO) Fetal Growth Charts based on fetal sex, gestational age, and weight.[4] Median and the interquartile range (IQR) for gestational days at delivery and birth weight were reported for unvaccinated, vaccinated, and boosted cohorts. A Fisher's Exact Test using R stats (version 4.1.1) was used to evaluate the difference in rates between the unvaccinated and all other cohorts as well as between boosted and vaccinated, but not boosted cohorts for categorical variables. A Mann-U Whitney test using Python scipy (version 1.4.1) was used to evaluate the differences between continuous variables.

**Supplemental Results**

*Efficacy of one mRNA dose, Ad26.COV2.S Johnson & Johnson, and COVID-19-induced immunity*

There was also a significantly lower rate of maternal COVID-19 infection in people who received one mRNA COVID-19 vaccine (n=104; 0.033, 95% CI[0.027, 0.039]; p<0.0001) or Ad26.COV2.S Johnson & Johnson (n=46; 0.034, 95% CI=[0.032, 0.036]; p<0.01), than in unvaccinated people (Supplemental Figure 5A). There was a significantly higher rate of maternal COVID-19 infection in unvaccinated people with COVID-19-induced immunity (n=67; 0.125, 95% CI=[0.100, 0.155]) compared to the unvaccinated population (p<0.0001). There is no difference in hospitalization rate during maternal COVID-19 infection with one mRNA COVID-19 vaccine dose (0.600, 95% CI=[0.487, 0.703], p=0.41) or Ad26.COV2.S Johnson & Johnson dose (0.559, 95% CI=[0.395, 0.711], p=1.0) compared to the unvaccinated cohort (Supplemental Figure 5B). There was a significantly higher rate of hospitalization during maternal COVID-19 infection in unvaccinated people with COVID-19-induced immunity (0.949, 95% CI=[0.861, 0.983]) compared to the unvaccinated population with no prior COVID-19 infection (p<0.001).

*Comparing the efficacy of mRNA-1273 Moderna and BNT162b2 Pfizer-BioNTech*

There were demographic differences between the type of COVID-19 vaccine (Moderna: n=9,981; Pfizer: n=16,811) received within the vaccinated cohort (Supplemental Table 10). Patients that received BNT162b2 Pfizer-BioNTech were significantly more likely to be: Black or Asian, Non-Hispanic, older, have lower pregravid BMI, have commercial insurance, non-smoker, non-illicit drug user, lower parity, lower gravidity, and live in urban areas, areas with lower socioeconomic vulnerability, lower household composition and disability vulnerability, and higher minority status and language vulnerability. There was no difference in fetal sex, mode of delivery, or housing type and transportation vulnerability level. There was no difference in the rates of chronic diabetes, chronic hypertension, gestational diabetes, or gestational hypertension amongst people receiving either vaccine. People that received BNT162b2 Pfizer-BioNTech had a significantly lower rate of preeclampsia, but no difference in the rate of severe preeclampsia compared to people that received mRNA-1273 Moderna.

Over a five-month period of pregnancy, people vaccinated with either mRNA-1273 Moderna (n=2,373; p<0.01) or BNT162b2 Pfizer-BioNTech (n=3,746; p<0.01) lower rates of COVID-19 infection compared to unvaccinated people matched on conception date (n=6,119; Supplemental Figure 6A). There was no difference in the infection rate between the two vaccines during this same period (p=0.30). 63% of mRNA fully vaccinated patients received BNT162b2 Pfizer-BioNTech (Supplemental Figure 6B). Breakthrough infections in people that were vaccinated with mRNA-1273 Moderna tended to occur significantly more days after achieving full vaccination status than those that received BNT162b2 Pfizer-BioNTech (p<0.01) indicating that they were protected from breakthrough infections for longer on average (Supplemental Figure 6C). People receiving either mRNA-1273 Moderna (0.037, 95% CI=[0.033, 0.041]; p<0.0001) or BNT162b2 Pfizer-BioNTech (0.032, 95% CI=[0.030, 0.035]; p<0.0001) were significantly less likely to have a maternal COVID-19 infection than unvaccinated people (0.07, 95% CI=[0.068, 0.072]; Supplemental Figure 6D, left panel). People that received BNT162b2 Pfizer-BioNTech were significantly less likely to have a maternal COVID-19 infection than those that received mRNA-1273 Moderna (p<0.05). There was no difference in the COVID-19-related hospitalization amongst people that were vaccinated with mRNA-1273 Moderna (0.526, 95% CI=[0.470, 0.580]; p=0.46) or BNT162b2 Pfizer-BioNTech (0.528, 95% CI=[0.482, 0.573]; p=0.76) and those that were unvaccinated (0.549, 95% CI=[0.526, 0.572]; Supplemental Figure 6D, right panel). There was also no difference in the COVID-19-related hospitalization in people that had been vaccinated with mRNA-1273 Moderna versus those vaccinated with BNT162b2 Pfizer-BioNTech (p=1.0).

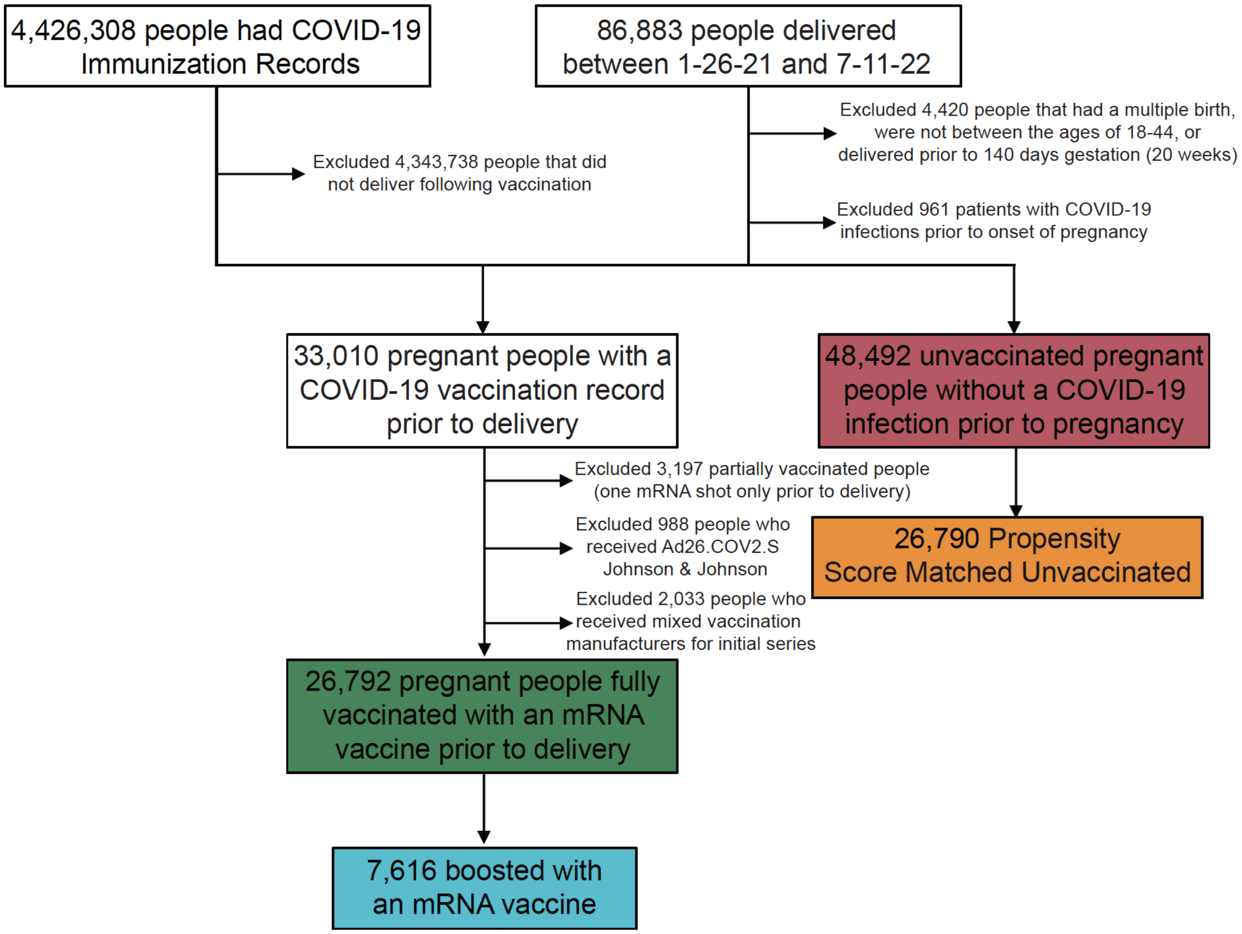


**Supplemental Figure 1. Flow diagram of cohort selection.**
COVID-19 mRNA vaccinated (two-dose vaccination of mRNA-1273 Moderna or BNT162b2 Pfizer-BioNTech) and unvaccinated (no COVID-19 vaccines received) cohorts were selected. These were defined using Providence St. Joseph Providence electronic health records, which include imported state vaccination records.

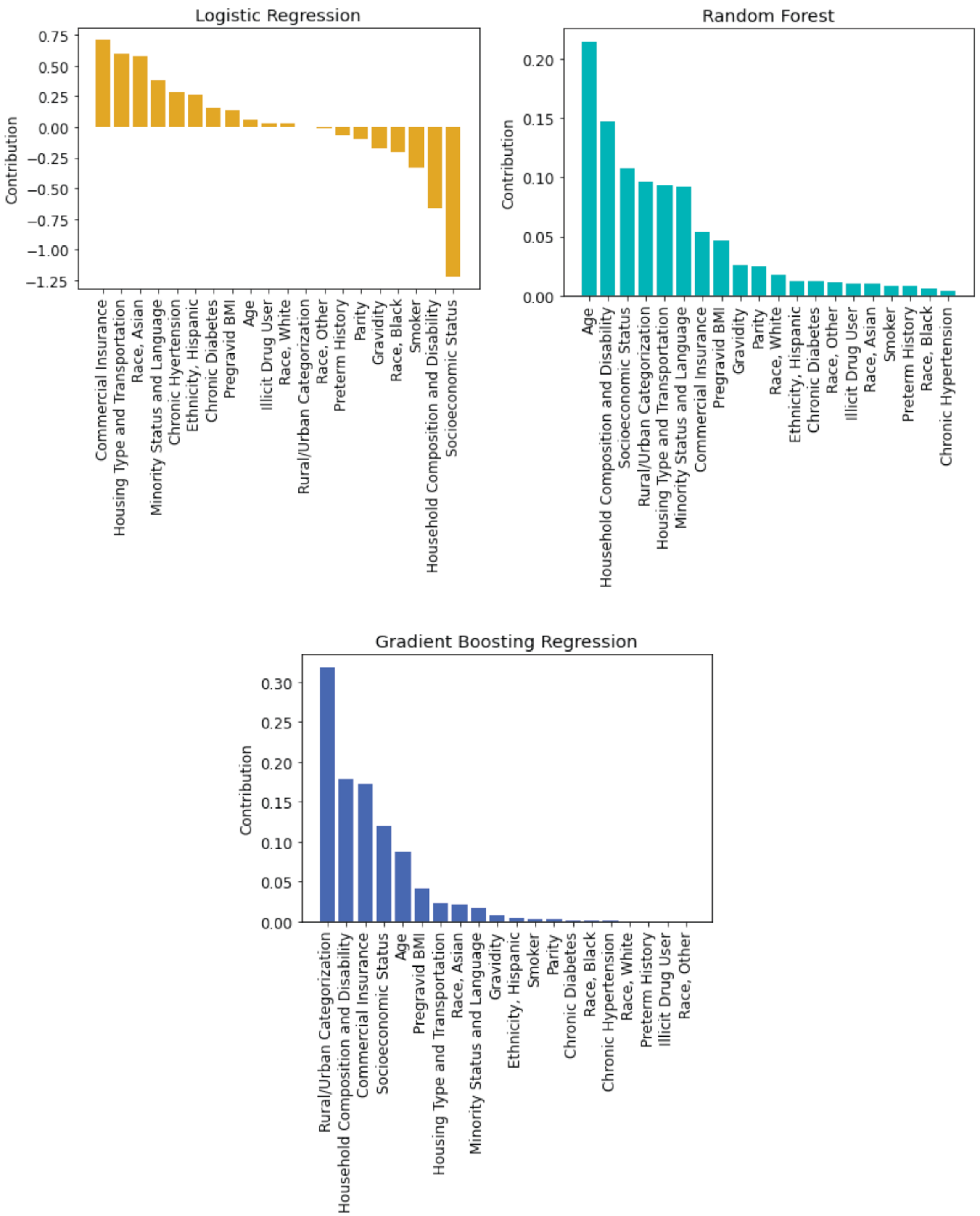


**Supplemental Figure 2. Contribution of features towards predicting vaccination status.**
Impurity-based feature importance evaluating the contribution of each of the 20 demographic, comorbidity, and geographical features for machine learning models predicting vaccination status at delivery. The importance of a feature is computed as the normalized total reduction of the criterion brought by that feature. The higher the value, the more important the feature. The models evaluated are logistic regression (gold; top left panel), random forest (teal; top right panel), and gradient boosting regression (blue; bottom panel).

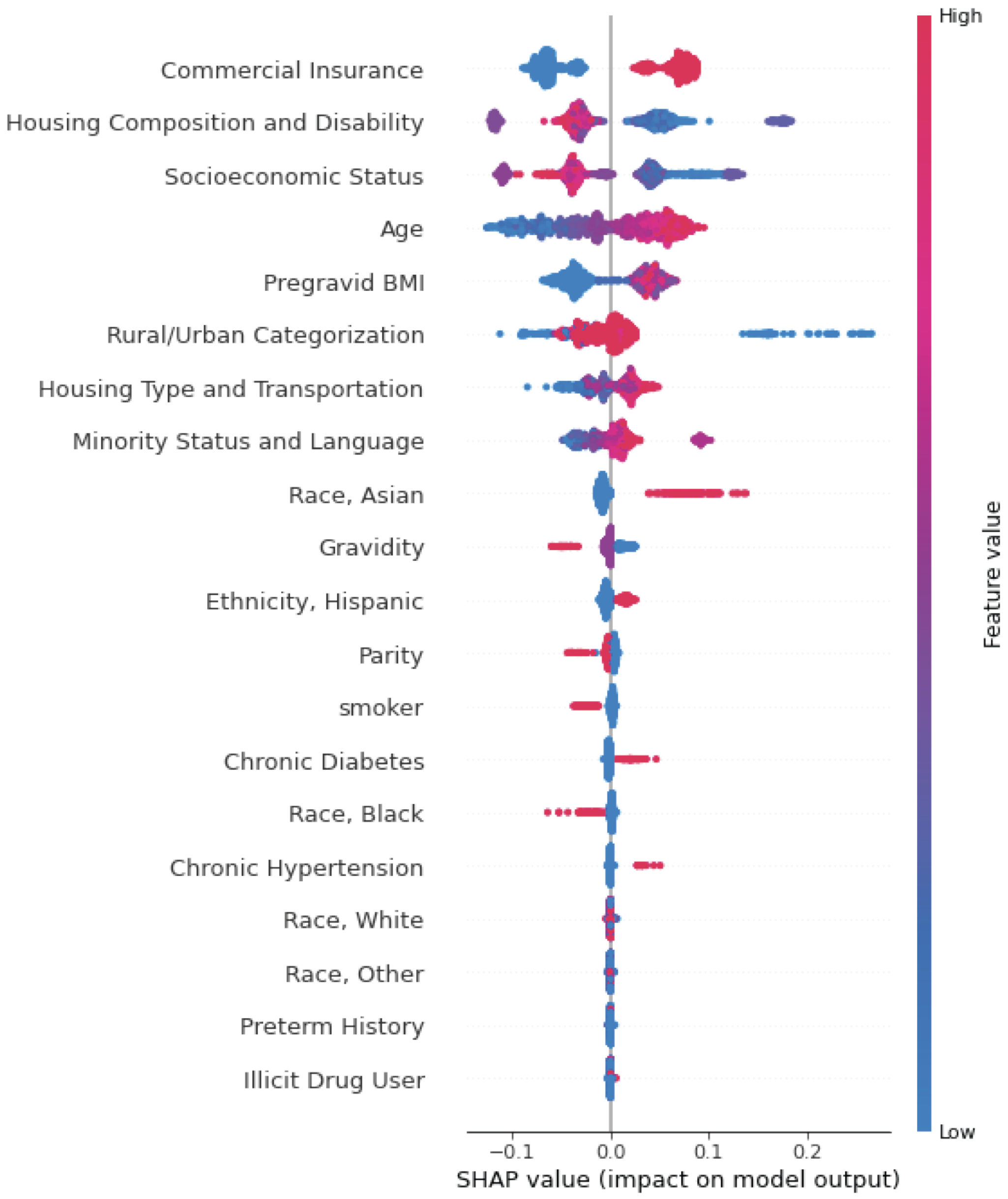


**Supplemental Figure 3. Shapley permutation explainer of the feature contribution towards classifying vaccination status.**

The contribution of all the features in the gradient boosting model towards classifying vaccination status at delivery as measured by the Shapley algorithm and reported as the SHAP value. This value is the average marginal contribution of a feature value across all permutations of features providing insight into the degree of influence of the feature on an individual's classified vaccination status at delivery. Each line represents a feature, and each dot represents a sample. The dot color represents the value of the feature for the sample, with red being a high value and blue being a low value for that feature across all samples. This evaluation was performed on 1000 patients randomly selected from the test set. SHAP=Shapley additive explanations.

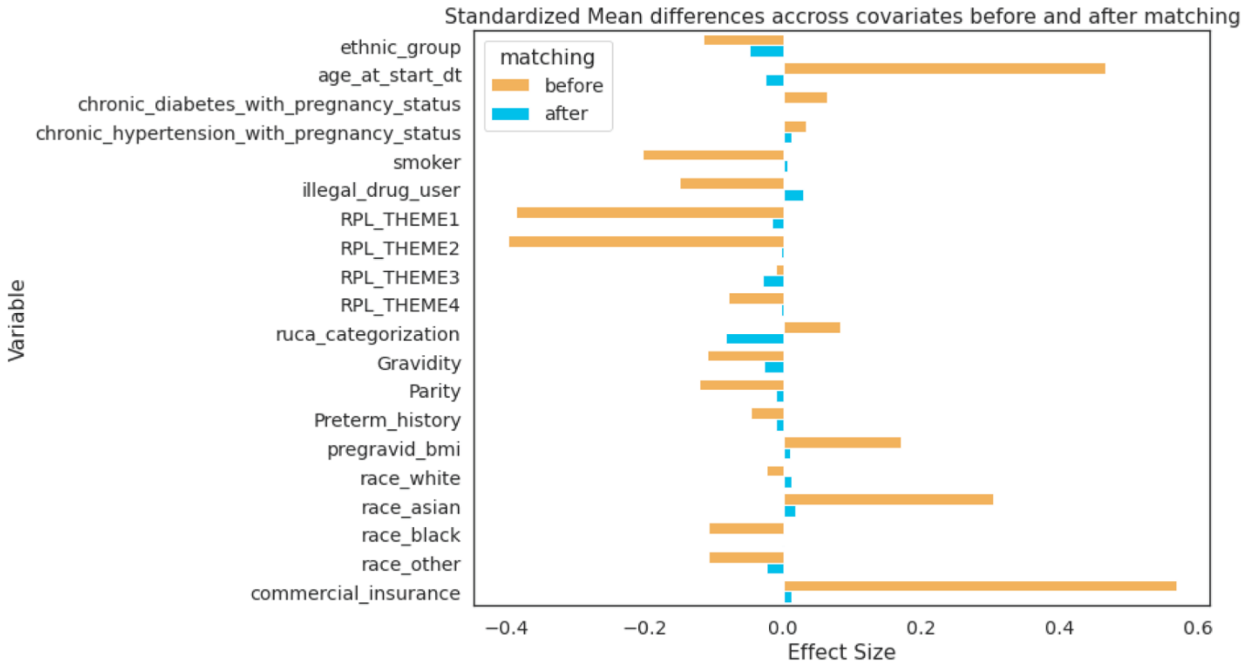


**Supplemental Figure 4. Propensity score matching reduces the differences in covariates between vaccinated and unvaccinated cohorts.**

Propensity score matching using nearest neighbors with replacement was performed using 20 covariates. The standardized mean difference between the vaccinated and the unvaccinated (before; orange) and unvaccinated matched (after; blue) is reported.

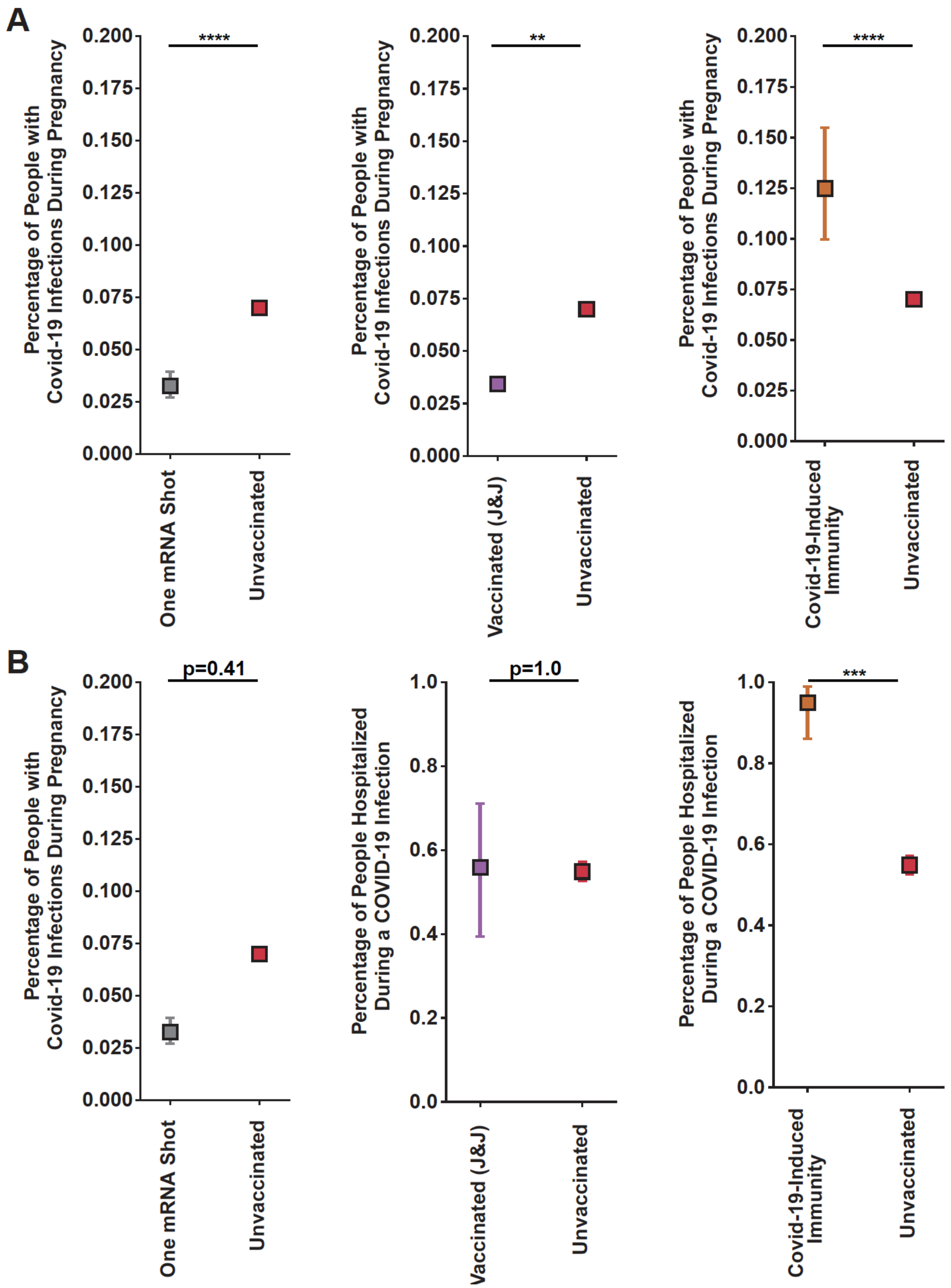


**Supplemental Figure 5. Rates of maternal COVID-19 infection and subsequent hospitalization in populations with immunity.**

**A:** Maternal COVID-19 infection rate with 95% CI in the unvaccinated population (red; n=48,492) compared to one mRNA dose (silver; n=3,194; left panel), Ad26.COV2.S Johnson & Johnson (purple; n=988; middle panel), or COVID-19-induced immunity (brown; n=537; right panel). **B:** Hospitalization rate during Omicron dominance (infection after 12/24/21) with 95% CI associated with maternal COVID-19 infection in the unvaccinated population (red; n=1,833) compared to one mRNA dose (silver; n=75; left panel); Ad26.COV2.S Johnson & Johnson (purple; n=34; center panel), or COVID-19-induced immunity (brown; n=59; right panel). 95% CI calculated by Wilson Score Interval and p-value by Fisher's Exact Test. ** p<0.01; *** p<0.001 **** p<0.0001.

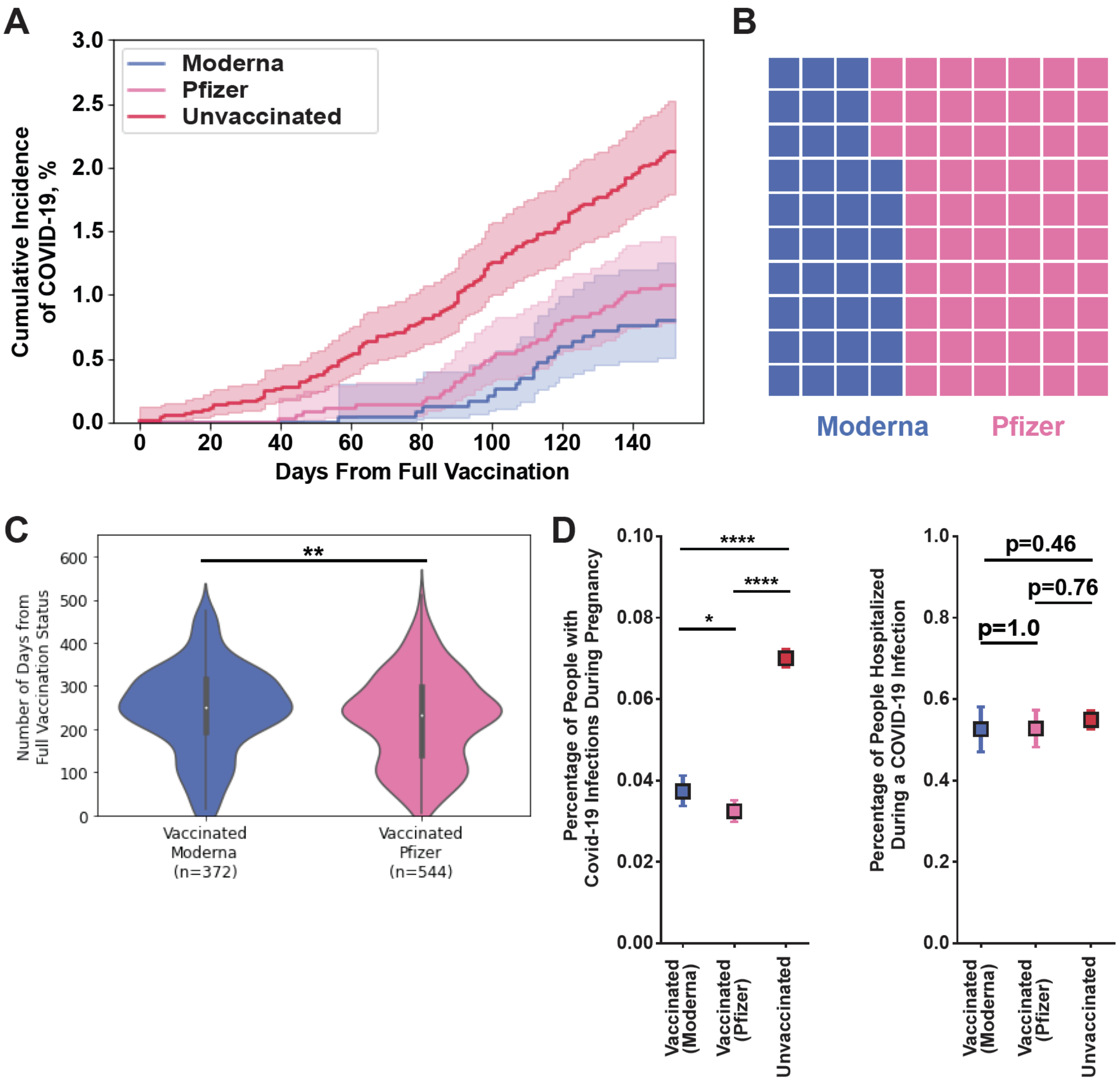


**Supplemental Figure 6. Effectiveness of mRNA-1273 Moderna and BNT162b2 Pfizer-BioNTech at preventing COVID-19 infections.**

**A:** Percentage of people plus the 95% CI that had COVID-19 infections over five months (152 days) of pregnancy in vaccinated individuals receiving mRNA-1273 Moderna (blue) or BNT162b2 Pfizer-BioNTech (pink) or those that were unvaccinated (red). T0 is at the day of full vaccination (2 weeks after the last dose of the initial vaccination series). Unvaccinated people were matched to vaccinated people on conception date and an unvaccinated person's T0 is the full vaccination date of the matched vaccinated person. p-values were calculated by a log rank test. Moderna: n=2,373; Pfizer n=3,746; Unvaccinated: n=6,119. Moderna vs Unvaccinated: $p<0.01$; Pfizer vs Unvaccinated: $p<0.01$; Moderna vs Pfizer $p=0.30$. **B:** Waffle plot of the percentage of the vaccinated cohort receiving Moderna (blue; n=9,981) or Pfizer (pink; n=16,811). **C:** Violin plot of the time from day of achieving full vaccination status to maternal breakthrough COVID-19 infection for individuals vaccinated with mRNA-1273 Moderna (blue; n=372) or BNT162b2 Pfizer-BioNTech (pink; n=554). The p-value was calculated by a Mann-U Whitney test. **D:** Percentage of people with 95% CI for people vaccinated with mRNA-1273 Moderna (blue), BNT162b2 Pfizer-BioNTech (pink), or unvaccinated (red) that had a maternal COVID-19 infection (left panel; Moderna n=9,981; Pfizer n=16,811; Unvaccinated n=48,492) or required hospitalization during a maternal COVID-19 infection during Omicron dominance (infection after 12/24/21; right panel; Moderna n=312; Pfizer n=455; Unvaccinated n=1,833). The 95% CI was calculated by Wilson Score Interval and the p-value was calculated by a Fisher's Exact Test. * $p<0.05$; ** $p<0.01$; **** $p<0.0001$.

**Supplemental Table 1. Vaccination status definitions.**
The definitions for different cohorts related to COVID-19 vaccination.

| Term | Definition |
|---|---|
| 1 mRNA Shot | A person who prior to delivery has received 1 mRNA COVID-19 vaccine dose: (Moderna or Pfizer) and has no recorded COVID-19 infection prior to the start of the current pregnancy. |
| Boosted | A person that prior to delivery received a third mRNA COVID-19 vaccine dose (Moderna or Pfizer), or booster, following completion of the initial series and has no recorded COVID-19 infection prior to the start of the current pregnancy. Booster dose is limited to mRNA dose, but does not have to match the brand (Moderna or Pfizer) of the initial vaccination series. |
| COVID-19-Induced Immunity | A person that has not received any COVID-19 vaccination dose regardless of brand prior to delivery and has previously had COVID-19 as reported by a PCR test in their EHR prior to the start of the current pregnancy. |
| Vaccinated | A person that has received at least two mRNA COVID-19 vaccine doses (Moderna or Pfizer) of the same brand at least two weeks prior to delivery and has no recorded COVID-19 infection prior to the start of the current pregnancy. |
| Vaccinated, but not boosted | A person that has received at least two mRNA COVID-19 vaccine dose (mRNA-1273 Moderna or BNT162b2 Pfizer-BioNTech) of the same brand more than five months prior to delivery, has not received a third booster dose prior to delivery, and has no recorded COVID-19 infection prior to the start of the current pregnancy. |
| Vaccinated, J&J | A person that has received at least one Ad26.COV2.S Johnson & Johnson (J&J) COVID-19 vaccine dose at least two weeks prior to delivery, has not received any other brand of COVID-19 dose, and has no recorded COVID-19 infection prior to the start of the current pregnancy. |
| Unvaccinated | A person that has not received any COVID-19 vaccination dose regardless of brand prior to delivery and has no recorded COVID-19 infection prior to the start of the current pregnancy. |
| Unvaccinated Matched | An unvaccinated person that was matched on several covariates to a vaccinated person using propensity score matching. |

**Supplemental Table 2. Feature definitions for demographic, birth characteristic, and geographical features.** The names and definitions of all the demographic and birth characteristics used to describe the cohort populations.

| Category | Feature | Definition |
|---|---|---|
| **Demographics** | Race | American Indian / Alaska Native, Asian, Black / African American, Native Hawaiian / Pacific Islander, Other White / Caucasian, or Unknown |
| | Ethnicity | Hispanic/Latino, Non-Hispanic/Latino, or Unknown |
| | Maternal Age | Maternal age (years) at start of pregnancy 18.0-44.9; reported as age ranges 18.0-24.9, 25.0-29.9, 30.0-34.9, 35.0-39.9, and 40.0-44.9 |
| | Pregravid BMI | Pregravid BMI (kg/m2); reported as categories Underweight (<18.5 BMI), Normal (18.5 - 24.9 BMI), Overweight ( 25.0 - 29.9 BMI), Obese (30.0 - 34.9 BMI), Severely Obese (>35.0 BMI), or Unknown |
| | Commercial Insurance | Health insurance provided and administered by non-governmental entities |
| | Smoker | History of smoking |
| | Illicit Drug Use | History of illicit drug use |
| | Preterm History | History of delivering prematurely |
| | Parity | Number of times a person has given birth to a fetus older than 24 weeks gestation prior to the current pregnancy; Nulliparity (0), Low Multiparity (1-3), or Grand Multipara (4+) |
| | Gravidity | Number of times a person has been pregnant regardless of the pregnancy outcome; Nulligravidity (0), Low Multigravidity (1-5), Grand Multigravidity (6+) |
| **Birth Characteristics** | Fetal Sex | Female, Male, or Unknown |
| | Mode of Delivery | C-Section, Vaginal, or Unknown |
| **Geographical Features** | Socioeconomic Status | CDC Social Vulnerability Index (SVI) Socioeconomic (RPL_THEME1) theme ranking; scores are 0-1 with 0 indicating low vulnerability on this theme; scores were mapped to patient U.S. Census Tract |
| | Housing Composition and Disability | CDC SVI Housing Composition & Disability (RPL_THEME2) theme ranking; scores are 0-1 with 0 indicating low vulnerability on this theme; scores were mapped to patient U.S. Census Tract |
| | Minority Status and Language | CDC SVI Minority Status & Language (RPL_THEME3) theme ranking; scores are 0-1 with 0 indicating low vulnerability on this theme; scores were mapped to patient U.S. Census Tract |
| | Housing Type and Transportation | CDC SVI Housing Type & Transportation (RPL_THEME4) theme ranking; scores are 0-1 with 0 indicating low vulnerability on this theme; scores were mapped to patient U.S. Census Tract |
| | Rural/Urban Categorization | U.S. Department of Agriculture Economic Research Service (USDA ERS) Rural-Urban Commuting Area (RUCA) codes; SecondaryRUCACode2010 (last updated in 2019) were mapped using patient U.S. Census Tract; Categorized as Metropolitan (> 4 score), Micropolitan (4 - 6 score), Small Town (7 - 9 score), Rural (10 - 99 score), or Unknown |

**Supplemental Table 3. Pregnancy-related conditions SNOMED definitions.**
The SNOMED Concept IDs used to define common comorbidities and pregnancy related conditions.

| Disease | SNOMED Concept ID | SNOMED Concept Name |
|---|---|---|
| **Diabetes mellitus (chronic)** | 199223000 | Diabetes mellitus during pregnancy, childbirth and the puerperium |
| | 199225007 | Diabetes mellitus during pregnancy - baby delivered |
| | 199227004 | Diabetes mellitus during pregnancy - baby not yet delivered |
| | 76751001 | Diabetes mellitus in mother complicating pregnancy, childbirth AND/OR puerperium |
| | 4783006 | Maternal diabetes mellitus with hypoglycemia affecting fetus OR newborn |
| | 10754881000119100 | Diabetes mellitus in mother complicating childbirth |
| | 199226008 | Diabetes mellitus in the puerperium - baby delivered during current episode of care |
| | 199228009 | Diabetes mellitus in the puerperium - baby delivered during previous episode of care |
| | 106281000119103 | Pre-existing diabetes mellitus in mother complicating childbirth |
| | 609563008 | Pre-existing diabetes mellitus in pregnancy |
| | 609564002 | Pre-existing type 1 diabetes mellitus in pregnancy |
| | 609567009 | Pre-existing type 2 diabetes mellitus in pregnancy |
| | 609566000 | Pregnancy and type 1 diabetes mellitus |
| | 237627000 | Pregnancy and type 2 diabetes mellitus |
| **Hypertension Disorder (chronic)** | 198941007 | Hypertension complicating pregnancy, childbirth, and the puerperium |
| | 541000119105 | Hypertension complicating pregnancy, childbirth, and the puerperium, antepartum |
| | 82771000119102 | Hypertension complicating pregnancy |
| | 37618003 | Chronic hypertension complicating AND/OR reason for care during pregnancy |
| | 24042004 | Chronic hypertension complicating AND/OR reason for care during puerperium |
| | 698640000 | Hypertension in the puerperium with pulmonary oedema |
| | 367390009 | Hypertension in the obstetric context |
| **Gestational Diabetes** | 393568003 | Gestational diabetes mellitus |
| | 11687002 | Gestational diabetes mellitus |
| | 40801000119106 | Gestational diabetes mellitus complicating pregnancy |
| | 10753491000119101 | Gestational diabetes mellitus in childbirth |
| | 75022004 | Gestational diabetes mellitus, class A>1< |
| | 46894009 | Gestational diabetes mellitus, class A>2< |
| | 40791000119105 | Postpartum gestational diabetes mellitus |
| **Gestational Hypertension** | 48194001 | Pregnancy-induced hypertension |
| | 237282002 | Impending eclampsia |
| | 288250001 | Maternal hypertension |
| | 698638005 | Pregnancy induced hypertension with pulmonary oedema |
| | 237281009 | Moderate proteinuric hypertension of pregnancy |
| | 307632004 | Non-proteinuric hypertension of pregnancy |
| | 237279007 | Transient hypertension of pregnancy |
| | 198965005 | Transient hypertension of pregnancy - delivered |
| | 198966006 | Transient hypertension of pregnancy - delivered with postnatal complication |
| | 198967002 | Transient hypertension of pregnancy - not delivered |
| | 198968007 | Transient hypertension of pregnancy with postnatal complication |
| | 15394000 | Toxaemia of pregnancy |
| | 40521000119100 | Postpartum pregnancy-induced hypertension |
| **Preeclampsia** | 398254007 | Pre-eclampsia |
| | 41114007 | Mild pre-eclampsia |
| | 765182005 | Pre-eclampsia in puerperium |
| | 46764007 | Severe pre-eclampsia |
| | 95605009 | Haemolysis-elevated liver enzymes-low platelet count syndrome |

| | | |
|---|---|---|
| | 198983002 | Severe pre-eclampsia - delivered |
| | 198984008 | Severe pre-eclampsia - delivered with postnatal complication |
| | 198985009 | Severe pre-eclampsia - not delivered |
| | 198986005 | Severe pre-eclampsia with postnatal complication |
| | 67359005 | Pre-eclampsia added to pre-existing hypertension |
| | 198997005 | Pre-eclampsia or eclampsia with pre-existing hypertension |
| | 198999008 | Pre-eclampsia or eclampsia with pre-existing hypertension - delivered |
| | 199000005 | Pre-eclampsia or eclampsia with pre-existing hypertension - delivered with postnatal complication |
| | 199002002 | Pre-eclampsia or eclampsia with pre-existing hypertension - not delivered |
| | 199003007 | Pre-eclampsia or eclampsia with pre-existing hypertension with postnatal complication |
| | 69909000 | Eclampsia added to pre-existing hypertension |
| **Severe Preeclampsia** | 198983002 | Severe pre-eclampsia - delivered (disorder) |
| | 198984008 | Severe pre-eclampsia - delivered with postnatal complication |
| | 198985009 | Severe pre-eclampsia - not delivered (disorder) |
| | 198986005 | Severe pre-eclampsia with postnatal complication |

**Supplemental Table 4. Feature definitions for models predicting vaccination status at delivery.**
The names of all the dependent variables and their definitions used in the Pearson correlations and supervised machine learning models.

| Feature | Definition |
|---|---|
| **Demographic** | |
| Race, Asian | Reported race: Non-Asian or Unknown (0), Asian (1) |
| Race, Black | Reported race: non-Black or Unknown (0), Black (1) |
| Race, Other | Reported race: Non-Other or Unknown (0), Other (1) |
| Race, White | Reported race: Non-White or unknown (0), White (1) |
| Ethnicity, Hispanic | Reported ethnicity: Non-Hispanic or Unknown (0) or Hispanic (1) |
| Maternal Age (Years) | Maternal age at start of pregnancy (years); continuous variable |
| Pregravid BMI (kg/m2) | Pregravid Body Mass Index (BMI; kg/m2); reported as categories Unknown (-1), Underweight (<18.5 BMI; 0), Normal (18.5 - 24.9 BMI; 1), Overweight (25.0 - 29.9 BMI; 2), Obese (30.0 - 34.9 BMI; 3), or Severely Obese (>35.0 BMI; 4) |
| Commercial Insurance | Health insurance provider: governmental health insurance, uninsured, or self-pay (0), or commercial insurance - Health insurance provided and administered by non-governmental entities |
| Smoker | Smoking status: no reported history of smoking (0), or reported history of smoking (1) |
| Illicit Drug User | Illicit drug status: no reported history of illicit drug use (0), or reported history of illicit drug use (1) |
| Preterm History | Previous preterm delivery: no reported history of premature delivery (0), or previously delivered prematurely prior to current pregnancy (1) |
| Parity | Number of times a person has given birth to a fetus older than 24 weeks of gestation prior to the current pregnancy: Nulliparity (0 births; 0), Low Multiparity (1 - 3 births; 1) or Grand Multipara (4+ births; 2) |
| Gravidity | Number of times a person has been pregnant regardless of the pregnancy outcome: Nulligravidity (0 pregnancies; 0), Low Multigravidity ( 1- 5 pregnancies; 1), Grand Multigravidity 6+ pregnancies; 2) |
| **Comorbidities** | |
| Chronic Diabetes | Chronic diabetes status: not diagnosed with chronic diabetes (0) or diagnosed with chronic diabetes (1) |
| Chronic Hypertension | Chronic hypertension status: not diagnosed with chronic hypertension (0) or diagnosed with chronic hypertension (1) |
| **Geographical Features** | |
| Socioeconomic Status | CDC Social Vulnerability Index (SVI) Socioeconomic theme: reported as a continuous variable [0-1] with 0 indicating a low vulnerability |
| Housing Composition and Disability | CDC SVI Housing Composition & Disability theme: reported as a continuous variable [0-1] with 0 indicating a low vulnerability |
| Minority Status and Language | CDC SVI Minority Status & Language theme: reported as a continuous variable [0-1] with 0 indicating a low vulnerability |
| Housing Type and Transportation | CDC SVI Housing Type & Transportation theme: reported as a continuous variable [0-1] with 0 indicating a low vulnerability |
| Rural/Urban Categorization | U.S. Department of Agriculture Economic Research Service (USDA ERS) Rural-Urban Commuting Area (RUCA) codes: reported as categories Unknown (-1), Rural (0), Small Town (1), Micropolitan (2), or Metropolitan (3) |

**Supplemental Table 5. Demographic, comorbidity, and geographical features association with vaccination status at delivery.**

Pearson's correlation coefficient and the corresponding p-values (two-tailed) for the correlation of vaccination status at delivery with the 20 features used in the predictive models.

| | Pearson Correlation | p-value |
|---|---|---|
| **Demographic** | | |
| Race, Asian | 0.14 | p<0.0001 |
| Race, Black | -0.05 | p<0.0001 |
| Race, Other | -0.05 | p<0.0001 |
| Race, White | -0.01 | p<0.01 |
| Ethnicity, Hispanic | -0.05 | p<0.0001 |
| Maternal Age (Years) | 0.22 | p<0.0001 |
| Pregravid BMI (kg/m2) | 0.08 | p<0.0001 |
| Commercial Insurance | 0.26 | p<0.0001 |
| Smoker | -0.10 | p<0.0001 |
| Illicit Drug User | -0.07 | p<0.0001 |
| Preterm History | -0.02 | p<0.0001 |
| Parity | -0.06 | p<0.0001 |
| Gravidity | -0.05 | p<0.0001 |
| **Comorbidities** | | |
| Chronic Diabetes | 0.03 | p<0.0001 |
| Chronic Hypertension | 0.01 | p<0.0001 |
| **Geographical Features** | | |
| Socioeconomic Status | -0.18 | p<0.0001 |
| Housing Composition and Disability | -0.19 | p<0.0001 |
| Minority Status and Language | -0.01 | p=0.16 |
| Housing Type and Transportation | -0.04 | p<0.0001 |
| Rural/Urban Categorization | 0.04 | p<0.0001 |

**Supplemental Table 6. Performance metrics of machine learning models predicting vaccination status at delivery.**

Performance metrics (mean absolute error, mean squared error, root mean squared error, area under the curve, and $R^2$) for the following models predicting the vaccination status (mRNA fully vaccinated or unvaccinated) at delivery: logistic regression, random forest, gradient regression, and gradient regression limited (top six features of the gradient boosting model only).

| | Logistic Regression | Random Forest | Boosting Regression | Boosting Regression Limited |
|---|---|---|---|---|
| Mean Absolute Error | 0.31 | 0.34 | 0.35 | 0.35 |
| Mean Squared Error | 0.31 | 0.18 | 0.17 | 0.17 |
| Root Mean Squared Error | 0.56 | 0.43 | 0.42 | 0.42 |
| Area Under the Curve | 0.63 | 0.77 | 0.80 | 0.79 |
| $R^2$ | -0.34 | 0.20 | 0.26 | 0.25 |

**Supplemental Table 7. Demographic, comorbidities, birth characteristics, and geographical features of unvaccinated matched people.**

Distribution of race, ethnicity, pregravid BMI, age, parity, gravidity, fetal sex, mode of delivery, and Rural/Urban Categorization for unvaccinated people that were propensity score matched to vaccinated people. Number of people who have commercial insurance, smoke, use illicit drugs, have previously delivered prematurely, or have common pregnancy-related comorbidities. Median and Interquartile Range (IQR) of CDC Social Vulnerability Index – socioeconomic status, household composition & disability, minority status and language, and housing type & transportation – for which a score [0,1] is assigned with 0 indicating low vulnerability on that theme. The p-values are comparing the vaccinated and unvaccinated matched pregnant populations and are calculated using a Fisher's Exact Test for categorical variables and Mann-U Whitney Test for continuous variables.

| | **No. (%)** | |
|---|---|---|
| | **Unvaccinated Matched n = 26,790** | **Vaccinated vs Unvaccinated Matched p-value** |
| **Demographics** | | |
| Race | | p<0.001 |
| American Indian or Alaska Native | 181 (0.7%) | |
| Asian | 3,908 (14.6%) | |
| Black | 843 (3.1%) | |
| Multiracial | 362 (1.4%) | |
| Native Hawaiian or Pacific Islander | 216 (0.8%) | |
| Other | 4,198 (15.7%) | |
| White | 15,735 (58.7%) | |
| Unknown | 1,347 (5.0%) | |
| Ethnicity | | p<0.001 |
| Hispanic or Latino | 6,786 (25.3%) | |
| Not Hispanic or Latino | 19,962 (74.5%) | |
| Unknown | 42 (1.6%) | |
| Maternal Age (Years) | | p<0.001 |
| 18-25 | 2,233 (8.3%) | |
| 25-30 | 5,459 (20.4%) | |
| 30-35 | 9,579 (35.8%) | |
| 35-40 | 7,459 (27.8%) | |
| 40-45 | 2,040 (7.6%) | |
| Pregravid BMI (kg/m2) | | p<0.001 |
| Underweight | 382 (1.4%) | |
| Normal | 5,765 (21.5%) | |
| Overweight | 4,431 (16.5%) | |
| Obese | 2,875 (10.7%) | |
| Severely Obese | 1,525 (5.7%) | |
| Unknown | 11,812 (44.1%) | |
| Commercial Insurance | 8,642 (32.3) | p=0.17 |
| Smoker | 1,164 (4.3%) | p=0.41 |
| Illicit Drug User | 1,692 (6.3%) | p<0.001 |
| Preterm History | 1,010 (3.8%) | p=0.23 |
| Parity | | p<0.001 |
| Nulliparity | 15,777 (58.9%) | |
| Low Multiparity | 10,478 (39.1%) | |
| Grand Multipara | 535 (2.0%) | |
| Gravidity | | p<0.001 |
| Nulligravidity | 8,712 (32.5%) | |
| Low Multigravidity | 16,753 (62.5%) | |
| Grand Multigravidity | 1,325 (4.9%) | |
| **Comorbidities** | | |
| Chronic Diabetes | 2,155 (7.8%) | p=0.99 |
| Chronic Hypertension | 483 (1.8%) | p=0.19 |
| Gestational Diabetes | 2,003 (7.5%) | p=0.74 |

| | | |
|---|---|---|
| Gestational Hypertension | 1,112 (4.0%) | p=0.30 |
| Preeclampsia | 681 (2.5%) | p<0.05 |
| Severe Preeclampsia | 15 (0.1%) | p=0.07 |
| **Birth Characteristics** | | |
| Fetal Sex | | p<0.05 |
| Female | 13,696 (51.1%) | |
| Male | 13046 (48.7%) | |
| Unknown | 50 (0.2%) | |
| Mode of Delivery | | p<0.05 |
| C-Section | 8,762 (32.7%) | |
| Vaginal | 17,974 (67.1%) | |
| Unknown | 54 (0.2%) | |
| **Geographical Features** | | |
| Rural/Urban Categorization | | p<0.001 |
| Rural | 228 (0.9%) | |
| Small Town | 275 (1.0%) | |
| Micropolitan | 692 (2.6%) | |
| Metropolitan | 22,702 (84.7%) | |
| Missing | 2,893 (10.8%) | |
| Socioeconomic Status | | |
| 1st Quintile (0.0 - 19.9%) | 9,852 (36.8%) | p<0.001 |
| 2nd Quintile (20.0 - 39.9%) | 6,773 (25.3%) | |
| 3rd Quintile (40.0 - 59.9%) | 5,251 (19.6%) | |
| 4th Quintile (60.0 - 79.9%) | 3,500 (13.1%) | |
| 5th Quintile (80.0% - 100.0%) | 1,414 (5.3%) | |
| Missing | 2,915 (10.9%) | |
| Household Composition and Disability | | p<0.001 |
| 1st Quintile (0.0 - 19.9%) | 12,684 (47.3%) | |
| 2nd Quintile (20.0 - 39.9%) | 6,280 (23.4%) | |
| 3rd Quintile (40.0 - 59.9%) | 4,002 (14.9%) | |
| 4th Quintile (60.0 - 79.9%) | 2,261 (8.4%) | |
| 5th Quintile (80.0% - 100.0%) | 1,563 (5.8%) | |
| Missing | 2,886 (10.8%) | |
| Minority Status and Language | | p<0.001 |
| 1st Quintile (0.0 - 19.9%) | 4,135 (15.4%) | |
| 2nd Quintile (20.0 - 39.9%) | 3,225 (12.0%) | |
| 3rd Quintile (40.0 - 59.9%) | 5,346 (20.0%) | |
| 4th Quintile (60.0 - 79.9%) | 8,719 (32.5%) | |
| 5th Quintile (80.0% - 100.0%) | 5,365 (20.0%) | |
| Missing | 2,886 (10.8%) | |
| Housing Type and Transportation | | p<0.001 |
| 1st Quintile (0.0 - 19.9%) | 5,760 (21.5%) | |
| 2nd Quintile (20.0 - 39.9%) | 4,920 (18.4%) | |
| 3rd Quintile (40.0 - 59.9%) | 4,433 (16.5%) | |
| 4th Quintile (60.0 - 79.9%) | 4,580 (17.1%) | |
| 5th Quintile (80.0% - 100.0%) | 7,097 (26.5%) | |
| Missing | 2,930 (10.9%) | |

**Supplemental Table 8. Dominant variant timing in the western United States.**

The time-period during which each variant was the dominant variant accounting for >50% of cases as part of the Center for Disease Control (CDC) genomic surveillance for SARS-CoV-2 in region 10 (Alaska, Idaho, Oregon, and Washington; CDC 2022).

| Dominant Variant | Start Date | End Date |
|---|---|---|
| Wild-Type | March 5 2020 | April 23, 2021 |
| Alpha | April 24, 2021 | July 2, 2021 |
| Delta | July 3, 2021 | December 24, 2021 |
| Omicron BA.1 | December 25, 2021 | March 25, 2022 |
| Omicron BA.2 | March 26, 2022 | May 27, 2022 |
| Omicron BA.2.12.1 | May 28, 2002 | July 1, 2022 |
| Omicron BA.5 | July 2, 2022 | July 11, 2022 |

**Supplemental Table 9. Additional outcomes related to maternal COVID-19 infection.**
Distribution of vaccinated (n=767), unvaccinated matched (n=437), or unvaccinated (n=1,833) people with maternal COVID-19 infections during Omicron dominance (infections after 12/24/21) for COVID-19 severity, supplemental oxygen use, and patient class. Median number and interquartile range (IQR) were reported for the total number and unique number of diagnoses made for patients during an active maternal COVID-19 infection. The p-values for vaccinated versus unvaccinated matched and vaccinated versus unvaccinated were calculated using a Fisher's Exact Test for categorical variables and a Mann-U Whitney Test for continuous variables.

| | **No. (%)** | **Median (IQR; [Min, Max])** | |
|---|---|---|---|
| | **Vaccinated n=767** | **Unvaccinated Matched n=437** | **Unvaccinated n=1,833** |
| COVID-19 Severity | | | |
| Mild | 692 (90.2%) | 379 (86.7%) | 1,672 (91.2%) |
| Moderate | 71 (9.3%) | 57 (13.0%) | 152 (8.3%) |
| Severe | 4 (0.5%) | 1 (0.2%) | 9 (0.5%) |
| p-value | | p=0.09 | p=0.70 |
| Max Oxygen Use | | | |
| None | 716 (93.4%) | 390 (89.2%) | 1,723 (94.0%) |
| Low-Flow Oxygen | 37 (6.1%) | 46 (10.5%) | 101 (5.5%) |
| High-Flow Oxygen | 2 (0.3%) | 1 (0.2%) | 5 (0.4%) |
| Ventilator | 2 (0.3%) | 0 (0.0%) | 4 (0.02%) |
| p-value | | p<0.05 | p=0.91 |
| Max Patient Class | | | |
| None | 85 (11.1%) | 86 (19.7%) | 265 (14.5%) |
| Outpatient | 278 (36.2%) | 137 (31.4%) | 562 (30.7%) |
| Emergency Care | 378 (49.3%) | 197 (45.1%) | 951 (51.9%) |
| Inpatient (Overnight Admittance) | 26 (3.4%) | 17 (3.9%) | 55 (3.0%) |
| p-value | | p<0.001 | p<0.05 |
| Number of Diagnoses, Unique | 3 (5; [0, 30]) | 2 (5; [0, 44]) | 2 (5; [0, 44]) |
| p-value | | p=0.12 | p=0.05 |
| Number of Diagnoses, Total | 3 (5; [0, 35]) | 2 (5; [0, 47]) | 2 (6; [0, 47]) |
| p-value | | p=0.07 | p<0.05 |
| Number of Medications, Inpatient | 9 (16; [0, 44]) | 9 (17 [0, 37]) | 10 (16; [0, 56]) |
| p-value | | p=0.43 | p=0.40 |
| Number of Medications, Outpatient | 2 (4; [0, 24]) | 2 (4 [0, 13]) | 2 (4; [0, 24]) |
| p-value | | p=0.15 | p=0.27 |
| Number of Medications, Total | 11 (18; [0, 61]) | 11 (19 [0, 44]) | 11 (18; [0, 95]) |
| p-value | | p=0.41 | p=0.41 |

**Supplemental Table 10. Demographic, comorbidities, birth characteristics, and geographical features of pregnant people vaccinated with Moderna vs Pfizer.**

Distribution of race, ethnicity, pregravid BMI, age, parity, gravidity, fetal sex, mode of delivery, and rural/urban categorization for people vaccinated by mRNA-1273 Moderna (n=9,981) or BNT162b2 Pfizer-BioNTech (n=16,811) at delivery. Number of people who have commercial insurance, smoke, use illicit drugs, have previously delivered prematurely, or have common pregnancy-related comorbidities. Median and Interquartile Range (IQR) of CDC Social Vulnerability Index – socioeconomic status, household composition & disability, minority status and language, and housing type & transportation – for which a score [0,1] is assigned with 0 indicating low vulnerability on that theme. The p-values are comparing the vaccinated and unvaccinated matched pregnant populations and are calculated using a Fisher's Exact Test for categorical variables and Mann-U Whitney Test for continuous variables.

| | **No. (%)** | | |
|---|---|---|---|
| | **Moderna**<br>**n = 9,981** | **Pfizer**<br>**n = 16,811** | **p-value** |
| **Demographics** | | | |
| Race | | | p<0.001 |
| American Indian or Alaska Native | 125 (1.3%) | 80 (0.5%) | |
| Asian | 1,374 (13.8%) | 2,777 (16.5%) | |
| Black | 283 (2.8%) | 573 (3.4%) | |
| Multiracial | 162 (1.6%) | 253 (1.5%) | |
| Native Hawaiian or Pacific Islander | 88 (0.9%) | 124 (0.7%) | |
| Other | 1,611 (16.4%) | 2,392 (14.2%) | |
| White | 5,962 (59.7%) | 9,924 (59.0%) | |
| Unknown | 376 (3.8%) | 688 (4.1%) | |
| Ethnicity | | | p<0.01 |
| Hispanic or Latino | 2,418 (24.2%) | 3,782 (37.9%) | |
| Not Hispanic or Latino | 7,555 (75.7%) | 13,007 (77.3%) | |
| Unknown | 8 (0.1%) | 22 (0.1%) | |
| Maternal Age (Years) | | | p<0.05 |
| 18-25 | 820 (8.2%) | 1,254 (7.5%) | |
| 25-30 | 2,050 (20.5%) | 3,300 (19.6%) | |
| 30-35 | 3,871 (38.8%) | 6,622 (39.4%) | |
| 35-40 | 2,693 (27.0%) | 4,641 (27.6%) | |
| 40-45 | 547 (5.5%) | 994 (5.9%) | |
| Pregravid BMI ( kg/m2) | | | p<0.001 |
| Underweight | 159 (1.6%) | 269 (1.6%) | |
| Normal | 2,417 (24.2%) | 4,308 (25.6%) | |
| Overweight | 1,732 (17.4%) | 2,958 (17.6%) | |
| Obese | 1,024 (10.3%) | 1,583 (9.4%) | |
| Severely Obese | 545 (5.5%) | 728 (4.3%) | |
| Unknown | 4,104 (41.1%) | 6,965 (41.4%) | |
| Commercial Insurance | 6,526 (65.4%) | 11,771 (70.0%) | p<0.0001 |
| Smoker | 497 (5.0%) | 707 (4.2%) | p<0.01 |
| Illicit Drug User | 765 (7.7%) | 1,125 (6.7%) | p<0.01 |
| Preterm History | 376 (3.8%) | 582 (3.5%) | p=0.20 |
| Parity | | | p<0.05 |
| Nulliparity | 5,762 (57.7%) | 10,001 (59.5%) | |
| Low Multiparity | 4,077 (40.8%) | 6,581 (39.1%) | |
| Grand Multipara | 142 (1.4%) | 229 (1.4%) | |
| Gravidity | | | p<0.05 |
| Nulligravidity | 3,176 (31.8%) | 5,635 (33.9%) | |
| Low Multigravidity | 6,405 (64.2%) | 10,549 (62.8%) | |
| Grand Multigravidity | 400 (4.0%) | 627 (3.7%) | |
| **Comorbidities** | | | |
| Chronic Diabetes | 829 (8.3%) | 1,325 (7.9%) | p=0.22 |
| Chronic Hypertension | 211 (2.1%) | 314 (1.9%) | p=0.18 |
| Gestational Diabetes | 763 (7.6%) | 1,220 (7.3%) | p=0.25 |

| | | | |
|---|---|---|---|
| Gestational Hypertension | 456 (4.6%) | 705 (4.2%) | p=0.15 |
| Preeclampsia | 324 (3.2%) | 450 (2.7%) | p<0.01 |
| Severe Preeclampsia | 13 (0.1%) | 15 (0.1%) | p=0.33 |
| **Birth Characteristics** | | | |
| Fetal Sex | | | p=0.39 |
| Female | 5,068 (50.8%) | 8,628 (51.3%) | |
| Male | 4,898 (49.1%) | 8,148 (48.5%) | |
| Unknown | 15 (0.2%) | 30 (0.2%) | |
| Mode of Delivery | | | p=0.69 |
| C-Section | 3,155 (31.6%) | 5,355 (31.9%) | |
| Vaginal | 6,806 (68.2%) | 11,425 (68.0%) | |
| Unknown | 20 (0.2%) | 31 (0.2%) | |
| **Geographical Features** | | | |
| Rural/Urban Categorization | | | p<0.001 |
| Rural | 126 (1.3%) | 78 (0.5%) | |
| Small Town | 91 (0.9%) | 114 (0.7%) | |
| Micropolitan | 409 (4.1%) | 354 (2.1%) | |
| Metropolitan | 8,028 (80.4%) | 13,912 (82.8%) | |
| Missing | 1,327 (13.3%) | 2,353 (13.9%) | |
| Socioeconomic Status | | | p<0.001 |
| 1st Quintile (0.0 - 19.9%) | 2,264 (22.7%) | 4,520 (26.9%) | |
| 2nd Quintile (20.0 - 39.9%) | 2,415 (24.2%) | 3,751 (22.3%) | |
| 3rd Quintile (40.0 - 59.9%) | 1,922 (19.3%) | 2,882 (17.1%) | |
| 4th Quintile (60.0 - 79.9%) | 1,381 (13.8%) | 2,183 (13.0%) | |
| 5th Quintile (80.0% - 100.0%) | 663 (6.6%) | 1,116 (6.6%) | |
| Missing | 1,336 (13.4%) | 2,359 (14.0%) | |
| Household Composition and Disability | | | p<0.001 |
| 1st Quintile (0.0 - 19.9%) | 3,214 (32.2%) | 5,958 (35.4%) | |
| 2nd Quintile (20.0 - 39.9%) | 2,102 (21.1%) | 3,542 (21.1%) | |
| 3rd Quintile (40.0 - 59.9%) | 1,500 (15.0%) | 2,504 (14.9%) | |
| 4th Quintile (60.0 - 79.9%) | 1,020 (10.2%) | 1,448 (8.6%) | |
| 5th Quintile (80.0% - 100.0%) | 820 (8.2%) | 1,012 (6.0%) | |
| Missing | 1,325 (13.3%) | 2,347 (14.0%) | |
| Minority Status and Language | | | p<0.001 |
| 1st Quintile (0.0 - 19.9%) | 576 (5.8%) | 854 (5.1%) | |
| 2nd Quintile (20.0 - 39.9%) | 1,360 (13.6%) | 2,029 (12.1%) | |
| 3rd Quintile (40.0 - 59.9%) | 1,859 (18.6%) | 3,138 (18.7%) | |
| 4th Quintile (60.0 - 79.9%) | 2,888 (28.9%) | 5,080 (30.2%) | |
| 5th Quintile (80.0% - 100.0%) | 1,973 (19.8%) | 3,363 (20.0%) | |
| Missing | 1,325 (13.3%) | 2,347 (14.0%) | |
| Housing Type and Transportation | | | p<0.001 |
| 1st Quintile (0.0 - 19.9%) | 980 (9.8%) | 1,749 (10.4%) | |
| 2nd Quintile (20.0 - 39.9%) | 1,705 (17.1%) | 2,993 (17.8%) | |
| 3rd Quintile (40.0 - 59.9%) | 1,680 (16.8%) | 2,673 (15.9%) | |
| 4th Quintile (60.0 - 79.9%) | 1,670 (16.7%) | 2,642 (15.7%) | |
| 5th Quintile (80.0% - 100.0%) | 2,610 (26.1%) | 4,395 (26.1%) | |
| Missing | 1,339 (13.4%) | 2,361 (14.0%) | |

**Supplemental Table 11. Additional birth outcomes by vaccination status.**
Number of people with low birth weight (<2,500 g) for vaccinated (n=26,792), unvaccinated matched (n=26,790), and unvaccinated (n=48,492) cohorts at delivery. Median number and interquartile range (IQR) were reported for the birth weight (g) and gestational days at delivery. The p-values were calculated using a Fisher's Exact Test for categorical variables and a Mann-U Whitney Test for continuous variables.

| | **No. (%)** | **Median (IQR)** | |
|---|---|---|---|
| | **Vaccinated n=26,792** | **Unvaccinated Matched n=26,790** | **Unvaccinated n=48,492** |
| Low Birth Weight (<2,500 g) | 1,524 (6.1%) | 1,638 (5.7%) | 3,107 (6.4%) |
| p-value | | p<0.05 | p<0.01 |
| Birth Weight (g) | 117.8 (22.8) | 118.2 (22.2) | 117.8 (22.9) |
| p-value | | p=0.15 | p=0.28 |
| Gestational Days at Delivery | 275 (11) | 274 (11) | 274 (12) |
| p-value | | p=0.24 | p<0.01 |

**Supplemental Table 12. Additional outcomes related to maternal COVID-19 infection.**
Distribution of two mRNA doses, but no booster (n=494) or boosted (three mRNA doses; n=437) people with maternal COVID-19 infections during Omicron dominance (infections after 12/24/21) for COVID-19 severity, supplemental oxygen use, and patient class. The number and percentage of patients that used supplemental oxygen or vasopressors during an active maternal COVID-19 infection are noted. Median number and interquartile range (IQR) were reported for the total number and unique number of diagnoses made for patients during an active maternal COVID-19 infection. The p-values for vaccinated versus unvaccinated matched and vaccinated versus unvaccinated were calculated using a Fisher's Exact Test for categorical variables and a Mann-U Whitney Test for continuous variables.

| | **No. (%)** | **Median (IQR; [Min, Max])** | |
|---|---|---|---|
| | **Vaccinated (2 mRNA doses) n=494** | **Boosted n=187** | **p-value** |
| COVID-19 Severity | | | p=0.27 |
| Mild | 451 (91.3%) | 165 (88.2%) | |
| Moderate | 41 (8.3%) | 20 (10.7%) | |
| Severe | 2 (0.4%) | 2 (0.1%) | |
| Max Oxygen Use | | | p=0.21 |
| None | 466 (94.3%) | 169 (90.4%) | |
| Low-Flow Oxygen | 26 (5.3%) | 16 (8.6%) | |
| High-Flow Oxygen | 1 (0.2%) | 1 (0.5%) | |
| Ventilator | 1 (0.2%) | 1 (0.5%) | |
| Max Patient Class | | | p=0.13 |
| None | 51 (10.3%) | 26 (13.9%) | |
| Outpatient | 173 (35.0%) | 77 (41.2%) | |
| Emergency Care | 254 (51.4%) | 80 (42.8%) | |
| Inpatient (Overnight Admittance) | 17 (3.4%) | 4 (2.1%) | |
| Supplemental Oxygen Use | 28 (5.7%) | 18 (9.6%) | p=0.09 |
| Vasopressor Use | 12 (2.5%) | 9 (4.8%) | p=0.13 |
| Number of Diagnoses, Unique | 3 (5; [0, 30]) | 3 (4; [0, 22]) | p=0.33 |
| Number of Diagnoses, Total | 3 (6; [0, 34]) | 3 (5; [0, 22]) | p=0.34 |
| Number of Medications, Inpatient | 7 (16; [0, 40]) | 10 (16; [0, 44]) | p=0.18 |
| Number of Medications, Outpatient | 2 (4; [0, 15]) | 3 (4; [0, 18]) | p<0.01 |
| Number of Medications, Total | 9 (17; [0, 61]) | 12 (18; [0, 50]) | p<0.05 |

**Supplemental Table 13. Additional birth outcomes by boosted status.**
Number of people with low birth weight (<2,500 g) for fully vaccinated, but not boosted (n=9,707) and boosted (n=7,558) at delivery. Median number and interquartile range (IQR) were reported for the birth weight (g) and gestational days at delivery. The p-values were calculated using a Fisher's Exact Test for categorical variables and a Mann-U Whitney Test for continuous variables.

| | **No. (%)** | **Median (IQR)** | |
|---|---|---|---|
| | **Vaccinated (2 mRNA doses) n=9,707** | **Boosted n=7,558** | **p-value** |
| Low Birth Weight (<2,500 g) | 673 (6.9%) | 431 (5.7%) | p<0.001 |
| Birth Weight (g) | 117.3 (23.5) | 118.2 (22.6) | p<0.05 |
| Gestational Days at Delivery | 274 (11.5) | 275 (11.0) | p=0.11 |
